# A distinct cross-reactive autoimmune response in multisystem inflammatory syndrome in children (MIS-C)

**DOI:** 10.1101/2023.05.26.23290373

**Authors:** Aaron Bodansky, Joseph J. Sabatino, Sara E. Vazquez, Janet Chou, Tanya Novak, Kristin L. Moffitt, Haleigh S. Miller, Andrew F. Kung, Elze Rackaityte, Colin R. Zamecnik, Jayant V. Rajan, Hannah Kortbawi, Caleigh Mandel-Brehm, Anthea Mitchell, Chung-Yu Wang, Aditi Saxena, Kelsey Zorn, David J.L. Yu, James Asaki, John V. Pluvinage, Michael R. Wilson, Laura L. Loftis, Charlotte V. Hobbs, Keiko M. Tarquinio, Michele Kong, Julie C. Fitzgerald, Paula S. Espinal, Tracie C. Walker, Stephanie P. Schwartz, Hillary Crandall, Katherine Irby, Mary Allen Staat, Courtney M. Rowan, Jennifer E. Schuster, Natasha B. Halasa, Shira J. Gertz, Elizabeth H. Mack, Aline B. Maddux, Natalie Z. Cvijanovich, Matt S. Zinter, Laura D. Zambrano, Angela P. Campbell, Adrienne G. Randolph, Mark S. Anderson, Joseph L. DeRisi, the Overcoming COVID-19 Network Study Group Investigators

## Abstract

Multisystem inflammatory syndrome in children (MIS-C) is a severe, post-infectious sequela of SARS-CoV-2 infection, yet the pathophysiological mechanism connecting the infection to the broad inflammatory syndrome remains unknown. Here we leveraged a large set of MIS-C patient samples (n=199) to identify a distinct set of host proteins that are differentially targeted by patient autoantibodies relative to matched controls. We identified an autoreactive epitope within SNX8, a protein expressed primarily in immune cells which regulates an antiviral pathway associated with MIS-C pathogenesis. In parallel, we also probed the SARS-CoV-2 proteome-wide MIS-C patient antibody response and found it to be differentially reactive to a distinct domain of the SARS-CoV-2 nucleocapsid (N) protein relative to controls. This viral N region and the mapped SNX8 epitope bear remarkable biochemical similarity. Furthermore, we find that many children with anti-SNX8 autoantibodies also have T-cells cross-reactive to both SNX8 and this distinct domain of the SARS-CoV-2 N protein. Together, these findings suggest that MIS-C patients develop a distinct immune response against the SARS-CoV-2 N protein that is associated with cross reactivity to the self-protein SNX8, demonstrating a link from the infection to the inflammatory syndrome.

## Introduction

Infection with severe acute respiratory syndrome coronavirus 2 (SARS-CoV-2) can lead to a broad spectrum of disease (*1–3*). The course of disease in children is often mild (*4–6*) but can become severe due to a variety of known (*7, 8*) and yet-to-be discovered factors. In rare cases children develop multisystem inflammatory syndrome in children (MIS-C), a post-infectious complication which often results in critical illness (*9–11*). The disease was hypothesized to be related to Kawasaki Disease, which presents similarly with prolonged fever, systemic inflammation, rash, conjunctivitis, and can be complicated by myocarditis and coronary artery aneurysms. In contrast to Kawasaki Disease, however, MIS-C is temporally associated after a SARS-CoV-2 infection, and more commonly includes shock, cardiac dysfunction, and multi-organ system involvement including gastrointestinal symptoms and hematologic findings, such as thrombocytopenia and lymphopenia (*13–17*). Through extensive characterization, MIS-C has been shown to have a distinctive inflammatory and cytokine signature, with evidence of altered innate and adaptive immunity as well as autoimmunity(*17–23*).

While the pathophysiological link between SARS-CoV-2 and MIS-C remains enigmatic, other autoimmune diseases are the consequence of exposure to novel antigens in the form of a virus or oncoprotein. For example, multiple sclerosis is associated with Epstein-Barr-Virus infection, and recent data suggests that B-cells and T-cells which are cross-reactive between a viral protein (EBNA1) and several host proteins may contribute to disease development(*24–26*). In addition, decades of paraneoplastic autoimmune encephalitis research, including anti-Hu, anti-Yo, anti-kelch-like protein 11, and many others, highlight the importance of autoreactive B-cells and T-cells working in concert to cause disease by targeting a shared intracellular antigen, and in certain cases a shared epitope (*28–35*).

Though cross-reactivity has not yet been identified in MIS-C, multiple autoantibodies have been reported (*20–22*), including those targeting interlueukin-1 receptor antagonist (IL-1Ra) (*35*). Distinctive T-cell signatures have also been reported in MIS-C, including an expansion of TRVB11-2 T-cells(*23, 36–39*) accompanied by autoimmune-associated B-cell expansions(*21*). Because the T-cell expansion is not monoclonal, many have speculated that a yet-to-be-identified superantigen is responsible. However, recent experiments suggest the expansion may instead by the result of activated tissue-resident T-cells(*41*), though the antigenic target of these T-cells remains unknown. Altered innate immune function has also been implicated in a subset of MIS-C cases, including inborn errors of immunity involving regulation of the mitochondrial antiviral-signaling (MAVS) protein pathway (*41*).

Here, children previously infected with SARS-CoV-2-with (n=199) and without (n=45) MIS-C were enrolled and comprehensively profiled for autoreactive antibodies as well as those targeting SARS-CoV-2. Differential autoreactivity highlighted an epitope motif which is shared by the viral N protein and the human SNX8 protein. SNX8 is expressed by immune cells across multiple tissues and modulates MAVS activity(*43*). This cross-reactive epitope motif is targeted by both B-cells and T-cells, suggesting that a subset of MIS-C may be triggered by molecular mimicry.

## Results

### MIS-C patients harbor a distinct set of autoreactivities

To explore the hypothesis that MIS-C is driven by an autoreactive process, the spectrum of autoantibody specificities present in children with MIS-C (n=199) and children who had asymptomatic or mild SARS-CoV-2 infection without MIS-C (within at least 5 weeks of enrollment) (n=45, hereafter referred to as “at-risk controls”) was captured using phage immunoprecipitation and sequencing (PhIP-Seq)(*44*). The PhIP-Seq technique has been extensively used to identify novel autoantigens in a wide range of diseases, and the specific 768,000 element human proteome-wide library used in these experiments has been used to define novel autoimmune syndromes and markers of disease for a variety of conditions (*29, 33, 44–46*). Given the inherently heterogeneous nature of antibody repertoires among individuals, the identification of disease-associated autoreactive antigens requires the use of large numbers of cases and controls (*45*). To minimize spurious hits, this study contains 199 MIS-C cases and 45 at-risk controls with recent SARS-CoV-2 infection, which represents substantially more MIS-C cases and controls than previously published (Figure 1A)(*20–22, 44*). Clinical characteristics of this cohort are described in Table 1.

**Figure 1:**
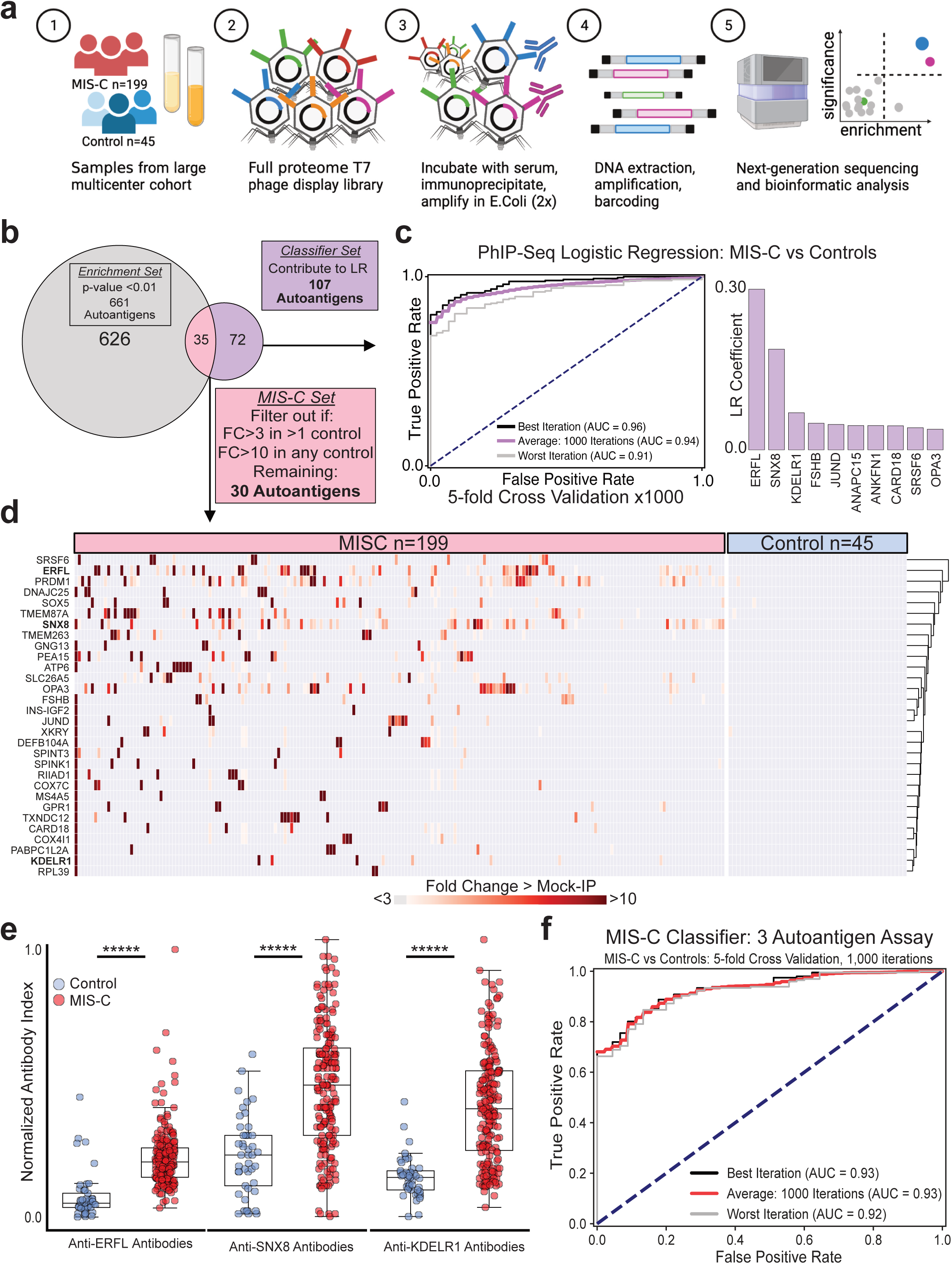
Autoantigens distinguish MIS-C from at-risk controls. (a) Design of PhIP-Seq experiment comparing MIS-C patients (n=199) and at-risk controls (n=45; children with SARS-CoV-2 infection at least 5 weeks prior to sample collection without symptoms of MIS-C). **(b)** Venn-diagram highlighting the number of autoantigens identified with statistically significant PhIP-Seq enrichment (‘*Enrichment set*’: Gray circle; p-value <0.01 on Kolmogorov-Smirnov (KS) test with Bonferroni FDR correction) and autoantigens identified which contribute to a logistic regression classifier of MIS-C relative to at-risk controls (*‘Classifier set*’: Purple circle). There are 35 autoantigens present in both the *classifer set* and *enrichment set* (Red; union of Venn Diagram) of which 30 are exclusive to MIS-C and referred to as the ‘*MIS-C Set’* (no two controls have low reactivity as defined by fold change signal over mean of Protein A/G beads only (FC > mock-IP) of 3 or greater, and no single control has high reactivity defined as FC > mock-IP greater than 10). **(c)** Receiver operating characteristic curve for the logistic regression classifier and barplot showing autoantigens with the top 10 logistic regression coefficients. **(d)** Hierarchically clustered (Pearson) heatmap showing the PhIP-Seq enrichment (FC > mock-IP) for the 30 autoantigens in the *MIS-C Set* in each MIS-C patient and each at-risk plasma control. (**e**) Radioligand binding assay (RLBA) values for 196 of the 199 individuals with MIS-C and each at-risk control for each of the top 3 autoantibodies identified by PhIP-Seq logistic regression. (**f**) Logistic regression receiver operating characteristic curve using input RLBA values to distinguish MIS-C from at-risk controls. For the figure, Mann-Whitney U testing was performed; *****p-value < 0.00001.

**Table 1:** Clinical characteristics of MIS-C and at-risk control cohorts.

|  | **MIS-C** | **Control** |
| --- | --- | --- |
| Number | 199 | 45 |
| Male sex (%) | 118 (59.3) | 24 (53.3) |
| Median age in years (median, IQR) | 10.9 (7.5 – 14.7) | 5.5 (2.2 – 11.1) |
| **Race and ethnicity (%)** |  |  |
| White, non-Hispanic | 63 (31.7) | 14 (31.1) |
| Black, non-Hispanic | 74 (37.2) | 5 (11.1) |
| Hispanic or Latino | 47 (23.6) | 15 (33.3) |
| Other race, non-Hispanic | 8 (4.0) | 2 (4.4) |
| Unknown | 7 (3.5) | 9 (20.0) |
| **Underlying conditions (%)** |  |  |
| None | 134 (67.3) | 36 (80) |
| Obesity* | 29 (15.2) | 8 (21.6) |
| Asthma | 22 (11.1) | 1 (2.2) |
| Cardiovascular | 4 (2.0) | 0 (0.0) |
| Immunocompromise | 0 (0.0) | 0 (0.0) |
| Autoimmune condition | 1 (0.5) † | 0 (0.0) |
| Malignancy | 1 (0.5) | 0 (0.0) |
| **Hospital Course (%)** |  |  |
| ICU admission | 170 (85.4) | 0 (0.0) |
| Shock requiring vasopressors | 90 (45.2) | 0 (0.0) |
| Mechanical ventilation | 38 (19.1) | 0 (0.0) |
| Extracorporeal membrane oxygenation (ECMO) | 8 (4.0) | 0 (0.0) |

\*Does not include n=8 MIS-C patients and n=11 controls under 2 years of age.
†Patient on chronic systemic steroids for eosinophilic esophagitis

For any given set of samples, PhIP-Seq has the potential to yield dozens to many thousands of differential enrichments of phage-displayed peptides. Here, logistic regression machine learning was used to first assess in an unbiased manner whether there existed a set of differentially enriched peptides that classify whether a sample is from an MIS-C patient or an at-risk control. This approach has been previously used with PhIP-Seq data to accurately classify Autoimmune Polyglandular Syndrome Type 1 (APS1) cases from controls (*45*). In this experiment, model input included the enrichments for all recovered peptide sequences derived from each sample. The training and classification were iterated 1,000 times using 5-fold cross validation, whereby 80% of the dataset is randomly chosen for training with the remaining 20% withheld for testing the classifier. Examination of the largest logistic regression coefficients associated with MIS-C disease revealed significant contributions to the classifier from peptides derived from the ERFL (ETS repressor factor like), SNX8 (Sorting nexin 8), and KDELR1 (KDEL endoplasmic reticulum protein retention receptor 1) coding sequences, followed by progressively smaller contributions from other genes. In all, 107 proteins had LR coefficients greater than zero, thus indicating greater autoantibody presence in MIS-C, and referred to here as the *classifier* set (Figure 1B). The receiver operating characteristic (ROC) curve for this analysis yielded a narrow range of values, with an average area under the curve (AUC) of 0.94 (Figure 1C).

In parallel, the Kolmogorov-Smirnov (KS) test was used to identify 661 statistically enriched autoreactivities (*statistically enriched set*: those with a p-value after adjustment for multiple test false discovery rates (FDR) less than 0.01). There is no agreed upon way to identify the most likely putative autoantibodies associated with disease among large peptidomic datasets. To avoid false positives, the intersection (n=35) of the *classifier* set and the *statistically enriched* set were considered further. Of these, peptides derived from 30 different genes also satisfied an additional set of conservative criteria, requiring that none be enriched (Fold-change over mock-IP (FC)>3) in more than a single control, or be enriched greater than 10-fold in any number of controls, hereafter referred to as the *MIS-C Set* (Figure 1B, Figure 1D). These 30 genes code for proteins with diverse function and tissue expression.

### Previously reported MIS-C autoantibodies

Approximately 34 candidate autoantigens have been previously reported as associated with MIS-C (*20–22, 44*). Of these, 33 were present in a similar proportion of MIS-C cases and at-risk controls in this cohort (Supplemental Figure 1A). Autoantibodies targeting the ubiquitously expressed ubiquitin protein ligase UBE3A were independently identified in this study as part of both the *classifier set* and the *statistically enriched set*, but we did not include them in the *MIS-C Set* given low positive signal in 2 controls.

Additionally, autoantibodies to the receptor antagonist IL-1Ra were previously reported in 13 of 21 (62%) MIS-C patients(*35*). In this cohort, anti-IL-1Ra antibodies were detected by PhIP-Seq (z-score > 6 over at-risk control) in 6 patient samples. To further examine immune reactivity to full length IL-1Ra, samples from 196 of the 199 patients in this study were used to immunoprecipitate S35 radiolabeled IL-1Ra (Methods). Positive immunoprecipitation of IL-1Ra (defined as greater than 3 standard deviations above mean of controls) was found in 39/196 (19.9%) patients with MIS-C, and the overall signal was significantly increased relative to at-risk controls (p-value <0.0001). However, many MIS-C patients were treated with intravenous immunoglobulin (IVIG), a blood product that has been shown to have autoantibodies(*48*). After removing samples of MIS-C patients who were treated with IVIG prior to sample collection (61 remaining), the difference between samples from MIS-C patients (5/61, 8.2%) and at-risk controls (1/45, 2.2%) was not significant (p-value = 0.30) (Supplemental Figure 1B).

### MIS-C autoantigens lack tissue specific associations with clinical phenotypes

Consistent with previous MIS-C reports(*10, 18*), this cohort was clinically heterogenous (Supplemental Table 1). To determine whether specific phenotypes, including myocarditis and the requirement of vasopressors, might be associated with specific autoantigens present in the *MIS-C Set*, protein tissue expression levels were assigned to each autoantigen (Human Protein Atlas; Proteinatlas.org), including amount of expression in cardiomyocytes and cardiac endothelium. The PhIP-Seq signal for MIS-C patients with a particular phenotype was compared to those MIS-C patients without the phenotype. Autoantigens with tissue specificity were not enriched in those MIS-C patients with phenotypes involving said tissue. Similarly, those autoantigens associated with myocarditis or vasopressor requirements did not correlate with increased cardiac expression (Supplemental Figure 2).

### Orthogonal validation of PhIP-Seq autoantigens

Peptides derived from ERFL, SNX8, and KDELR1 carried the largest logistic regression coefficients in the MIS-C classifier. The PhIP-Seq results were orthogonally confirmed by immunoprecipitation of S35 radiolabeled full-length ERFL, SNX8, and KDELR1 (radioligand binding assay (RLBA), see Methods) with patient (196 of the 199 samples used for PhIP-Seq) and at-risk control (all 45) samples. Relative to at-risk controls, samples from MIS-C patients significantly enriched each of the three target proteins (p-value <0.00000001 for ERFL, SNX8, and KDELR1), consistent with the PhIP-Seq assay (Figure 1E). Using only the radiolabeled immunoprecipitation data for these three proteins, MIS-C could be confidently classified (using 5-fold cross validation, iterated 1,000 times) from at-risk control sera with a ROC AUC of 0.93, suggesting potential for molecular diagnostic purposes (Figure 1F).

In this cohort, IVIG was administered to 137 of the 199 patients with MIS-C prior to sample collection and was absent from all 45 at-risk controls. The autoreactivity to the ERFL, SNX8, and KDELR1 proteins from the 62 MIS-C patients who had not been treated with IVIG prior to sample collection were compared to the at-risk controls (Supplemental Figure 3A). In contrast to IL-1Ra, the differential enrichment of these three proteins remained significant (p-value <0.0001, <0.00007, and <0.00011 respectively) suggesting that autoreactivity to ERFL, SNX8, and KDELR1 proteins was not confounded by IVIG treatment (Supplemental Figure 3B).

### Independent MIS-C cohort validation

To further test the validity of these findings, an independent validation cohort consisting of samples from 24 different MIS-C patients and 29 children severely ill with acute COVID-19 infection was acquired (Supplemental Table 2). Using the RLBA assay with full length SNX8, ERFL, and KDELR1 proteins, all three target proteins were significantly enriched relative to both the at-risk controls (p-value = 0.00023, 0.00004, and 0.00003 for ERFL, SNX8, and KDELR1, respectively) and the severe acute COVID-19 patients (p-value = 0.007, 0.008, 0.0012 for ERFL, SNX8, and KDELR1, respectively) (Supplemental Figure 4A). Using a logistic regression classifier that was trained on the original cohort, the ROC AUC in the validation cohort for classification of MIS-C from at-risk controls was 0.84, and from severe acute pediatric COVID-19 was 0.78 (Supplemental Figure 4B), suggesting that autoreactivity to these three proteins is a significant feature of MIS-C separable not only from SARS-CoV-2 exposure itself, but also from severe acute pediatric COVID-19.

### MIS-C autoantibodies target a single epitope within SNX8 protein

SNX8 is a protein of 465 amino acids that belongs to a family of sorting nexins involved in endocytosis, endosomal sorting, and signaling(*49*). Publicly available expression data (Human Protein Atlas; Proteinatlas.org) shows SNX8 is widely expressed across a variety of tissues including the brain, heart, gastrointestinal tract, kidneys, and skin, with highest expression in undifferentiated cells and immune cells. Previous work has associated SNX8 with host defense against bacteria, DNA viruses, and RNA viruses (*43, 50, 51*). ERFL is a poorly characterized protein of 354 amino acids. A survey of single cell RNA sequencing data (Human Protein Atlas; Proteinatlas.org) suggests enrichment in plasma cells, B-cells, and T-cells in some tissues. Using a Spearman correlation in principal component analysis (PCA) space based on tissue RNA sequencing data (Human Protein Atlas; Proteinatlas.org) SNX8 has the second closest expression pattern to ERFL, with a correlation of 0.81. KDELR1 is a 212 amino acid ER-Golgi transport protein essential to lymphocyte development with low tissue expression specificity. All three proteins are predicted or known to be intracellular, suggesting that putative autoantibodies targeting these proteins are unlikely to be sufficient for disease pathology on their own. However, autoantibodies targeting intracellular antigens are often accompanied by a T-cell which is autoreactive to the protein from which that antigen was derived, and which targets cell types expressing the protein. Of these proteins, we selected SNX8 for further investigation given its enrichment in immune cells, as well as its putative role in regulating the MAVS pathway in response to RNA viral infection, a pathway recently implicated in MIS-C pathology(*41*).

Full length SNX8 is represented in this PhIP-Seq library by 19 overlapping 49mer peptides. For all but one patient sample, the peptide fragment spanning amino acid positions 25 to 73 was the most enriched in the PhIP-Seq assay (Figure 2A), suggesting a common autoreactive site. To narrow the portion of this peptide to a smaller fragment, a sequential alanine scan was performed (Figure 2B; Methods). Using samples from six individuals with MIS-C, the critical region for immunoreactivity was determined to span 9 amino acids, positions 51 through 59 (PSRMQMPQG). Using the wild-type 49 amino acid peptide and the version with the critical region mutated to alanines, 182 of the 199 MIS-C patients and all 45 controls were assessed for immunoreactivity using a split-luciferase binding assay (Methods; SLBA). We found that samples from 31 of 182 (17.0%) patients with MIS-C immunoprecipitated the wildtype fragment. Of these, 29 (93.5%) failed to immunoprecipitate the mutated peptide, suggesting a common shared autoreactive epitope among nearly all of the MIS-C patients with anti-SNX8 antibodies. In contrast, only a single sample (out of 45) from the at-risk control cohort immunoprecipitated the wildtype fragment in this assay (Supplemental Figure 5).

**Figure 2:**
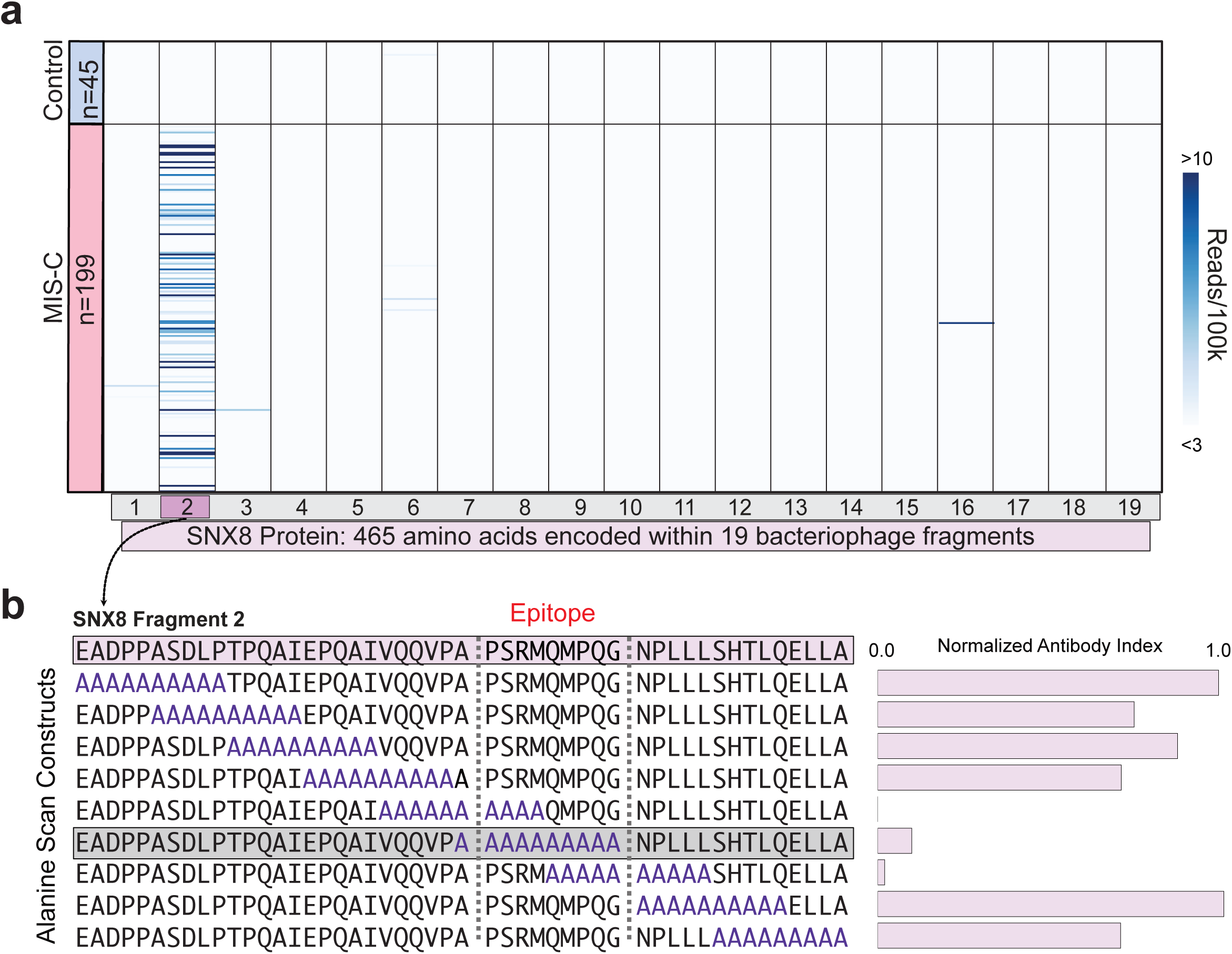
Autoantibodies in MIS-C patients target a single epitope within SNX8. (**a**) PhIP-Seq signal (reads per 100k) for each MIS-C patient (n=199) and each at-risk control (n=45) across each of the 19 bacteriophage encoded peptide fragments which together tile the full length SNX8 protein. (**b**) SLBA enrichments (normalized antibody indices) for each sequential alanine mutagenesis construct. Constructs were designed with 10 amino-acid alanine windows shifted by 5 amino-acids until the entire immunodominant SNX8 region (SNX8 Fragment 2) was scanned. Values are averages of 6 separate MIS-C patients. Identified autoantibody epitope bounded by vertical gray dotted lines.

### MIS-C patients have an altered antibody response to the SARS-CoV-2 N protein

To evaluate whether differences exist in the humoral immune response to SARS-CoV-2 infection in patients with MIS-C relative to at-risk controls, we repeated PhIP-Seq with 181 of the original 199 MIS-C patients and all 45 of the at-risk controls using a previously validated library specific for SARS-CoV-2(*52*). To discover whether certain fragments were differentially enriched by either MIS-C or at-risk controls, the enrichment of each phage encoded SARS-CoV-2 peptide (38 amino acids each) in each of the MIS-C patients and each at-risk control was normalized to 48 pre-COVID healthy controls. Three nearly adjacent peptides derived from the SARS-CoV-2 N protein were found to have significant differences in enrichment (KS-test, p< 0.000001 for each of the three peptides). The first peptide, spanning amino acids 77-114, was significantly enriched in the at-risk controls (representing the typical serological response in children), whereas the next two fragments, spanning amino acids 134-190, were significantly enriched by MIS-C patients (Figure 3A, B). This differentially reactive domain of the SARS-CoV-2 N protein was termed the MIS-C associated domain of SARS-CoV-2 (MADS). The PhIP-Seq results were orthogonally confirmed using the SLBA assay to measure the amount of MADS peptide immunoprecipitated with samples from 16 individuals, including 11 MIS-C patients and 5 at-risk controls (Figure 3C). To more precisely map the critical reactive region of the differentially enriched N protein peptides by MIS-C samples, peptides featuring a sliding window of 10 alanine residues were used as the immunoprecipitation substrate for the SLBA assay, run in parallel with the SNX8 alanine scanning peptides for three MIS-C sera samples (Figure 3D). Remarkably, the critical regions identified here in both SNX8 and SARS-CoV-2 N protein possessed significant similarity and can be represented by the regular expression [ML]Q[ML]PQG (Figure 3E).

**Figure 3:**
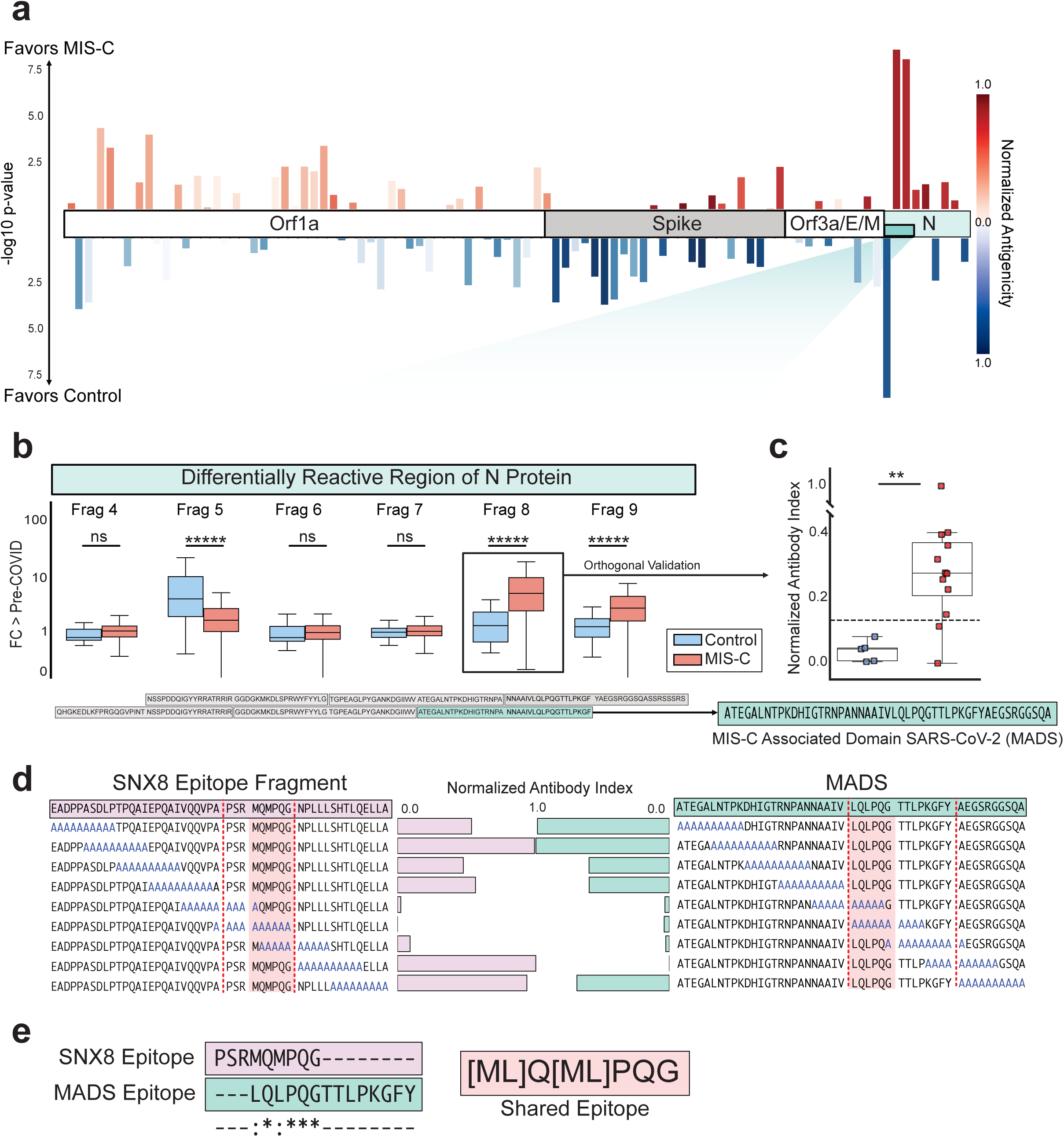
MIS-C patient antibodies preferentially target distinct region of SARS-CoV-2 N protein. (**a**) Relative PhIP-Seq signal (fold-change over mean of 48 pre-COVID controls (FC > Pre-COVID)) in MIS-C patients (n=181) and at-risk controls (n=45) using custom phage display library expressing entire SARS-CoV-2 proteome to different regions of SARS-CoV-2. Only regions with mean antibody signal >1.5 fold above pre-COVID controls shown. Antigenicity (sum of mean FC > pre-COVID in MIS-C and at-risk controls) represented by darker shades. Length of bars represents statistical difference in signal between MIS-C and at-risk controls to a particular region (-log 10 of Kolmogorov-Smirnov test p-values), with upward deflections representing enrichment in MIS-C versus at-risk controls, and downward deflections representing less signal in MIS-C. (**b**) Barplots showing PhIP-Seq signal (FC > Pre-COVID) across the specific region of the SARS-CoV-2 N protein with most divergent response in MIS-C samples relative to at-risk controls. Amino acid sequence of region with highest relative enrichment in MIS-C highlighted in green, and referred to as the MIS-C associated domain of SARS-CoV-2 (MADS). **(c)** Stripplots showing MADS SLBA enrichments (normalized antibody indices) in MIS-C patients (n=11) relative to at-risk controls (n=5). (**d**) SLBA signal (normalized antibody indices) for full sequential alanine mutagenesis scans within the same 3 individuals for SNX8 (left) and MADS (right) identification of epitopes to each (bounded by red dotted vertical lines). (**e)** Multiple sequence alignment of SNX8 and MADS epitopes (ClustalOmega; asterisk=identical amino acid; colon=strongly similar properties with Gonnet PAM 250 matrix score >0.5) with amino acid sequence for similarity region shown (orange). For the figure, Kolmogorov-Smirnov testing was used to compare PhIP-Seq signal, and Mann-Whitney U testing was used to compare SLBA signal; ns p-value > 0.05, **p-value <0.01, *****p-value < 0.00001.

### MIS-C patients have significantly increased SNX8 autoreactive T-cells

In other autoimmune diseases, autoantibodies are often generated against intracellular targets, yet the final effectors of cellular destruction are autoreactive T-cells(*31, 52*). Given evidence that certain subsets of MIS-C are HLA-associated(*36, 37*), and that SNX8 is known to be an intracellular protein, we hypothesized that MIS-C patients with anti-SNX8 antibodies may, in addition to possessing SNX8 autoreactive B-cells, also possess autoreactive T-cells targeting SNX8 expressing cells. To test this hypothesis, T-cells from 9 MIS-C patients (8 from SNX8 autoantibody positive patients and 1 who was SNX8 autoantibody negative) and 10 at-risk controls (chosen randomly) were exposed to a pool of 15-mer peptides with 11 amino acid overlaps that collectively tiled full length human SNX8 protein. T-cell activation was measured by an activation induced marker (AIM) assay, which quantifies upregulation of three cell activation markers: OX40, CD69 and CD137 (Figure 4A)(*53–55*). The percent of T-cells which activated in response to SNX8 protein was significantly higher in MIS-C than controls (p-value <0.001) Using a positive cutoff of 3 standard deviations above the mean of the controls, 7 of the 9 MIS-C (78%) patients were deemed positive for SNX8 autoreactive T-cells, while no controls met these criteria (Figure 4B). With respect to CD8+ and CD4+ subgroups specifically, there was increased signal in MIS-C relative to controls which did not meet significance (p=0.071 and p=0.058, respectively) (Supplemental Figure 6). The MIS-C patient who was seronegative for the SNX8 autoantibody was also negative for SNX8 autoreactive T-cells.

**Figure 4:**
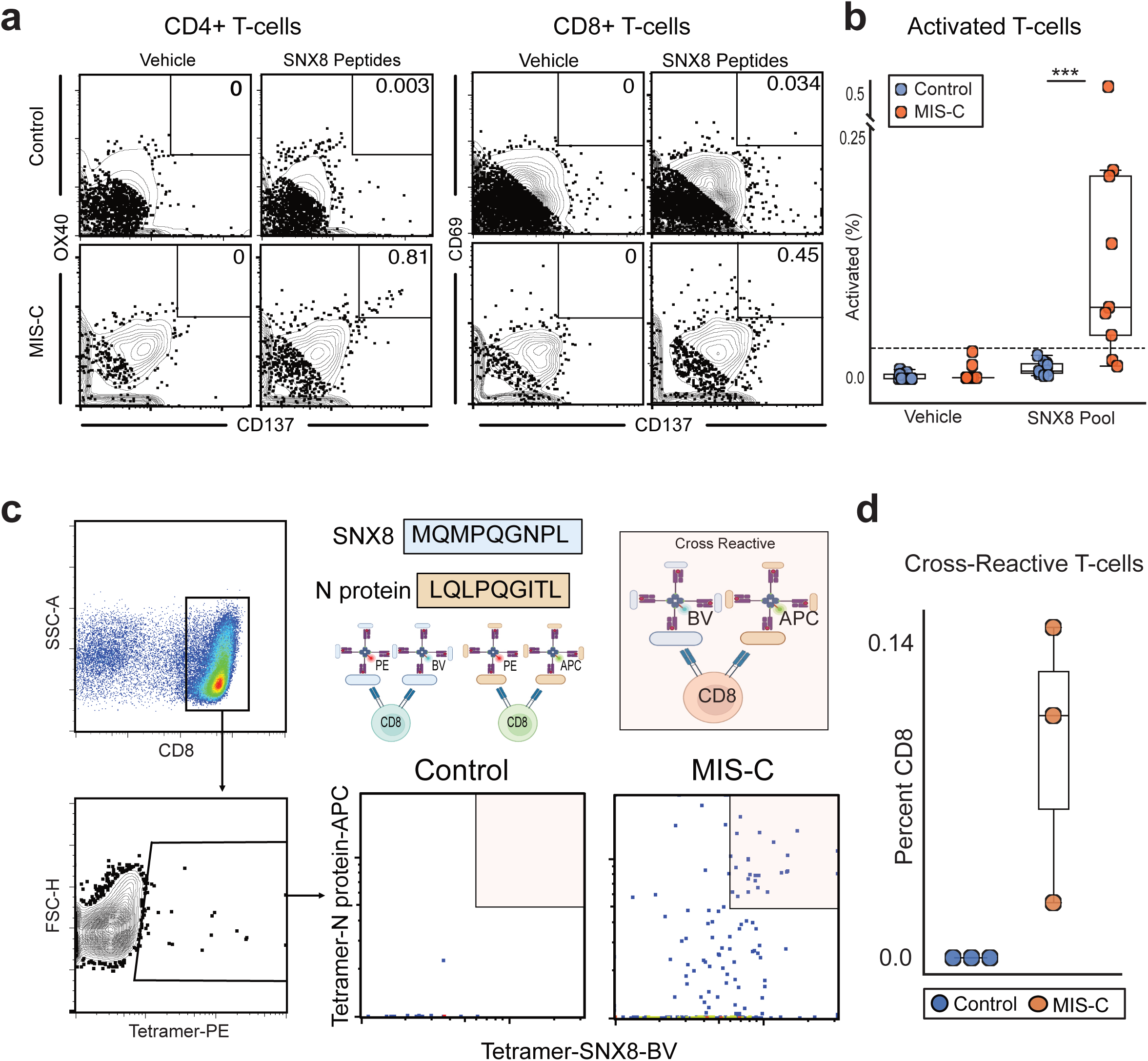
MIS-C patients harbor T-cells autoreactive to SNX8 which cross-react with SARS-CoV-2. (**a**) Representative activation induced marker (AIM) assay flow cytometry gating strategy measuring percent of CD4+ T-cells which activate in response to SNX8 protein (CD137+/OX40+) and percent of CD8+ T-cells which activate (CD137+/CD69+). **(b)** Barplot showing distribution of T-cells which activate in response to either vehicle (culture media + 0.2% DMSO) or SNX8 peptide pool (SNX8 peptide + culture media + 0.2% DMSO). **(c)** Gating strategy used to identify CD8+ T-cells which bound to SNX8 epitope and/or MADS N protein epitope (CD8+ T-cells positive for PE). Representative MIS-C patient and control showing each CD8+ T-cell which bound to any tetramer (PE+) and the relative binding of that T-cell to both the SNX8 epitope (BV421+) and the MADS N protein epitope (APC+) identifying cross-reactive T-cells (PE+/ APC+/BV421+). (**d**) Barplot showing percentage of CD8+ T-cells which are cross-reactive to both SNX8 and MADS in MIS-C and controls (3 MIS-C patients and 3 at-risk controls). For the figure, Mann-Whitney U testing was performed; ***p-value <0.001.

### HLA type A*02 is more likely to present the shared epitope

MIS-C has been associated with HLA types A*02, B*35, and C*04(*36, 37*). The Immune Epitope Database and Analysis Resource (IEDB.org(*56*)) was used to rank the MHC1 peptide presentation likelihoods for both SNX8 and SARS-CoV-2 N protein with respect to the MIS-C associated HLA alleles. The distribution of predicted MHC I binding scores for N protein and SNX8 fragments matching the [ML]Q[ML]PQG regular expression relative to fragments lacking a match was compared. For HLA type A*02, predicted MHC I binding was significantly higher (p<0.0001 for N protein and p=0.001 for SNX8) for fragments containing the putative autoreactive motif. There was no statistical difference for HLA B*35 and C*04 predictions (Supplemental Figure 7). Of note, of the 7 MIS-C patients with SNX8 autoreactive T-cells, 4 of the patients had HLA-typing performed, and all 4 were HLA-A*02 positive (Supplemental Figure 6).

### Identification of MIS-C T-cells cross-reactive to the similarity regions of SNX8 and N protein

Given the prediction that MIS-C associated HLA types preferentially display peptides containing the similarity regions for both SNX8 and the SARS-CoV-2 N protein, we sought to determine whether cross-reactive T-cells were present, and if they were specific to MIS-C. We therefore stimulated PBMCs from 3 MIS-C patients and 3 at-risk controls with peptides from either the SNX8 similarity region [MQMPQGNPL] or the SARS-CoV-2 N protein similarity region [LQLPQGITL] for 7 days to enrich for CD8+ T-cells reactive to these epitopes. We then built differently labeled MHCI tetramers loaded with either the SNX8 peptide [MQMPQGNPL] or the SARS-CoV-2 peptide [LQLPQGITL] (Figure 4C) and measured binding to T-cells.

Each of the three MIS-C patients had multiple CD8+ T-cells which bound to both the SNX8 peptide and the SARS-CoV-2 peptide. None of the three at-risk controls had any cross-reactive CD8+ T-cells. The percentage of CD8+ T-cells binding to each of the individual epitopes was also measured (Figure 4D). The 3 MIS-C patients had multiple T-cells which bound to the N protein peptide, SNX8 peptide, and both peptides (cross-reactive).

### SNX8 peptide with similarity region is sufficient to activate T-cells

Because MIS-C patients preferentially harbor T-cells which activate in response to a peptide pool containing fragments spanning the entire SNX8 protein and have T-cells which are cross-reactive T-cells between similarity regions of SNX8 and the N-protein, the similarity region of SNX8 alone was tested for its ability to activate patient T-cells. A pool of 20 10mer peptides with 9 amino acid overlaps centered on the target motif from SNX8 (collectively spanning amino acids 44 through 72) was used to stimulate PBMCs from two patients with MIS-C and 4 at-risk controls. Both of the MIS-C patients had activation of T-cells and none of the 4 controls had T-cell activation (Supplemental Figure 8).

### RNA expression profile of SNX8 during SARS-CoV-2 infection

As previously discussed, SNX8 is widely expressed across many tissue types, but most highly expressed by immune cells, consistent with its known function defending against RNA viruses via recruitment of MAVS(*43*). To further investigate the potential impact of combined B-cell and T-cell autoimmunity to SNX8 in the setting of SARS-CoV-2 infection, we analyzed SNX8 expression from single-cell sequencing of PBMC samples from patients with severe, mild, or asymptomatic COVID-19 infection, influenza infection, and uninfected healthy controls (*57*). In the setting of SARS-CoV-2 infection, SNX8 has highest mean expression in classical and non-classical monocytes and B-cells (Supplemental Figure 9A and Supplemental Figure 9B). SNX8 has higher expression in individuals with SARS-CoV-2 infections compared to uninfected (Supplemental Figure 9C). Within myeloid lineage cells, SNX8 expression correlates with MAVS expression, and OAS1 and OAS2 (two known regulators of the MAVS pathway specifically implicated in MIS-C pathogenesis(*41*)) expression (Supplemental Figure 9D). In contrast to MAVS, SNX8 expression is inversely correlated to severity of SARS-CoV-2 infection, with highest expression in asymptomatic individuals. This follows a similar pattern to OAS1 and OAS2. However, unlike OAS1, OAS2, and MAVS, SNX8 is preferentially expressed in SARS-CoV-2 infections relative to influenza virus infection (Supplemental Figure 9E).

## Discussion

The SARS-CoV-2 pandemic largely spared children from severe disease. One rare but notable exception is MIS-C, an enigmatic and life-threatening syndrome. Children with MIS-C have increased circulating inflammatory markers, dysregulated innate and adaptive immunity, and evidence of autoimmunity (*17–23, 37, 58*). Previous studies have surfaced numerous associations, but have failed to identify a direct mechanistic link between SARS-CoV-2 and MIS-C. Among those previously reported associated autoantibodies (*20–22, 44*), only autoantibodies targeting UBE3A and IL1RA were significantly enriched in this study, though anti-UBE3A antibodies were found in some controls and an association with anti-IL1RA antibodies were confounded by IVIG treatment.

In this study, 199 MIS-C patient samples and 45 pediatric at-risk controls were analyzed using a customized human and SARS-CoV-2 proteome PhIP-seq library. In the human library, the results reveal an autoreactive signature that distinguishes cases from controls, driven by enrichment of peptides deriving from three proteins, including SNX8, which is expressed by immune cells across multiple tissues. For SARS-CoV-2, differential enrichment between cases and controls was dominated by peptides derived from the N protein. Fine epitope mapping of both SNX8 and N protein identified a sequence that is highly similar, represented by the regular expression [ML]Q[ML]PQG. Furthermore, CD8+ T-cells were found to be cross-reactive for this same epitope in MIS-C cases, but not controls. These cross-reactive cells may contribute to immune dysregulation through the inappropriate engagement of immune cell lineages expressing SNX8. Intriguingly, we found evidence for sharing of the [ML]Q[ML]PQG epitope between both B-cells and T-cells in our analysis and further study will be needed to understand this apparent convergence. Importantly, there is broad evidence that in many autoimmune diseases autoantibodies can target intracellular antigens, but pathology is thought to be driven by an autoreactive T-cell response(*31, 52*).

These findings may help to clarify and connect several important known aspects of MIS-C pathophysiology. Specifically, these results draw parallels to other diseases in which exposure to a new antigen leads to autoimmunity, such as paraneoplastic autoimmune disease or cross-reactive viral epitopes between EBV and host proteins in multiple sclerosis(*24–26, 31*). An expansion of TRVB11-2 T-cells in MIS-C has been shown(*23, 36–39*), and while a superantigen has been postulated, it has yet to be identified. Recent studies have shown that tissue resident T-cells exhibit site-specific expansions(*59*), and that MIS-C patients have a distinct increase in activated circulating T-cells with a tissue-resident phenotype(*41*), leading to speculation that these T-cells may be driving the TRVB11-2 expansion. Although SNX8 is a relatively understudied protein, it has been linked to the function and activity of MAVS(43). Importantly, dysregulation of the MAVS antiviral pathway, by inborn errors of immunity, has been shown to underly certain cases of MIS-C(*41*). The most straightforward connection linking MIS-C to SNX8 may be through an inappropriate autoimmune response against tissues with significant MAVS pathway expression, but further investigation will be needed to test this potential link.

In addition to helping contextualize previous findings, these results are the first to directly link the initial SARS-CoV-2 infection and the subsequent development of MIS-C. We propose that MIS-C may be the results of multiple uncommon events converging. The initial insult is likely the formation of a combined B-cell and T-cell response that preferentially targets a particular motif within the N protein of SARS-CoV-2. A subset of individuals may then develop a cross-reactive B-cell and T-cell response to the self-protein SNX8. Interestingly, this cross-reactive motif has strong binding characteristics for the MIS-C associated HLA-A*02(*37*), further indicating that this may be an important risk factor in the development of MIS-C.

These results describe a subset of MIS-C (at least 17% positive for antibodies against SNX8), indicating that other mechanisms likely exist. Antibodies against ERFL are present in many children with MIS-C who do not have autoreactivity to SNX8, and ERFL has a highly similar tissue RNA expression profile to SNX8 (2nd most similar among all known proteins; Human Protein Atlas; Proteinatlas.org). If autoreactive T-cells to ERFL indeed also exist, they would be predicted to engage a nearly identical set of cells and tissues. It is important to also consider that MIS-C prevalence has rapidly decreased as an increasing number of children have developed immunity through vaccination and natural SARS-CoV-2 infection. We speculate that perhaps this could be related to the strong deviation of the anti-SARS CoV-2 immune response away from the critical region of the N-protein that we have identified, to other major epitopes such as those in the Spike protein through vaccination and past infection. Alternatively, ongoing evolution of SARS-CoV-2 may include mutations in the N protein that could affect subsequent antigen processing and presentation.

MIS-C is complex and more work is required to fully unravel the etiology. Why the differential adaptive immune response outlined above occurs in children with MIS-C and not others remains somewhat unclear. However, the results of this study, and specifically the development of combined cross-reactive B-cells and T-cells, bear similarity to other notable examples of molecular mimicry. Future work is needed to examine the mechanisms by which a cross-reactive epitope forces a break in tolerance, which may point toward better diagnostics and therapeutic interventions for many diseases with similar underlying mechanisms.

## Methods

### Patients

Patients were recruited through the prospectively enrolling multicenter Overcoming COVID-19 and Taking on COVID-19 Together study in the United States. The study was approved by the central Boston Children’s Hospital Institutional Review Board (IRB) and reviewed by IRBs of participating sites with CDC IRB reliance. A total of 292 patients were enrolled into 1 of the following independent cohorts between June 1, 2020 and September 9, 2021: 223 patients hospitalized with MIS-C (199 in the primary discovery cohort, 24 in a separate subsequent validation cohort), 29 patients hospitalized for COVID-19 in either an intensive care or step-down unit (referred to as severe acute COVID-19 in this study), and 45 outpatients (referred to as “at-risk controls” in this study) post-SARS-CoV-2 infections associated with mild or no symptoms. The demographic and clinical data are summarized in Table I, Supplemental Table 1, and Supplemental Table 2. The 2020 US Centers for Disease Control and Prevention case definition was used to define MIS-C(*60*). All patients with MIS-C had positive SARS-CoV-2 serology results and/or positive SARS-CoV-2 test results by reverse transcriptase quantitative PCR. All patients with severe COVID-19 or outpatient SARS-CoV-2 infections had a positive antigen test or nucleic acid amplification test SARS-CoV-2. For outpatients, samples were collected from 36 to 190 days after the positive test (median, 70 days after positive test; interquartile range, 56-81 days). For use as controls in the SARS-CoV-2 specific PhIP-Seq, plasma from 48 healthy, pre-COVID-19 controls were obtained as deidentified samples from the New York Blood Center. These samples were part of retention tubes collected at the time of blood donations from volunteer donors who provided informed consent for their samples to be used for research.

### DNA oligomers for split luciferase binding assays (SLBAs)

DNA coding for the desired peptides for use in split luciferase binding assays were inserted into split luciferase constructs containing a terminal HiBiT tag and synthesized (Twist Biosciences) as DNA oligomers and verified by Twist Biosciences prior to shipment. Constructs were amplified by PCR using the 5’-AAGCAGAGCTCGTTTAGTGAACCGTCAGA-3’ and 5’-GGCCGGCCGTTTAAACGCTGATCTT-3’ primer pair.

For SNX8, the oligomers coded for the following sequences:

EADPPASDLPTPQAIEPQAIVQQVPAPSRMQMPQGNPLLLSHTLQELLA AAAAAAAAAATPQAIEPQAIVQQVPAPSRMQMPQGNPLLLSHTLQELLA EADPPAAAAAAAAAAEPQAIVQQVPAPSRMQMPQGNPLLLSHTLQELLA EADPPASDLPAAAAAAAAAAVQQVPAPSRMQMPQGNPLLLSHTLQELLA EADPPASDLPTPQAIAAAAAAAAAAAPSRMQMPQGNPLLLSHTLQELLA EADPPASDLPTPQAIEPQAIAAAAAAAAAAQMPQGNPLLLSHTLQELLA EADPPASDLPTPQAIEPQAIVQQVPAAAAAAAAAANPLLLSHTLQELLA EADPPASDLPTPQAIEPQAIVQQVPAPSRMAAAAAAAAAASHTLQELLA EADPPASDLPTPQAIEPQAIVQQVPAPSRMQMPQGAAAAAAAAAAELLA EADPPASDLPTPQAIEPQAIVQQVPAPSRMQMPQGNPLLLAAAAAAAAA

For SARS-CoV-2 nucleocapsid, the oligomers coded for the following sequences:

ATEGALNTPKDHIGTRNPANNAAIVLQLPQGTTLPKGFYAEGSRGGSQA AAAAAAAAAADHIGTRNPANNAAIVLQLPQGTTLPKGFYAEGSRGGSQA ATEGAAAAAAAAAAARNPANNAAIVLQLPQGTTLPKGFYAEGSRGGSQA ATEGALNTPKAAAAAAAAAANAAIVLQLPQGTTLPKGFYAEGSRGGSQA ATEGALNTPKDHIGTAAAAAAAAAALQLPQGTTLPKGFYAEGSRGGSQA ATEGALNTPKDHIGTRNPANAAAAAAAAAAGTTLPKGFYAEGSRGGSQA ATEGALNTPKDHIGTRNPANNAAIVAAAAAAAAAAKGFYAEGSRGGSQA ATEGALNTPKDHIGTRNPANNAAIVLQLPQAAAAAAAAAAEGSRGGSQA ATEGALNTPKDHIGTRNPANNAAIVLQLPQGTTLPAAAAAAAAAAGSQA ATEGALNTPKDHIGTRNPANNAAIVLQLPQGTTLPKGFYAAAAAAAAAA

### DNA plasmids for radioligand binding assays (RLBAs)

For radioligand binding assays, DNA expression plasmids under control of a T7 promoter and with a terminal Myc-DDK tag for the desired protein were utilized. For ERFL, a custom plasmid was ordered from Twist Bioscience in which a Myc-DDK-tagged full length ERFL sequence under a T7 promoter was inserted into the pTwist Kan High Copy Vector (Twist Bioscience). Twist Bioscience verified a sequence perfect clone by next generation sequencing prior to shipment. Upon receipt, the plasmid was sequence verified by Primordium Labs. For SNX8, a plasmid containing Myc-DDK-tagged full length human SNX8 under a T7 promoter was ordered from Origene (RC205847) and was sequence verified by Primordium Labs upon receipt. For KDELR1, a plasmid containing Myc-DDK-tagged full length human KDELR1 under a T7 promoter was ordered from Origene (RC205880), and was sequence verified by Primordium Labs upon receipt. For IL1RN, a plasmid containing Myc-DDK-tagged full length human IL1RN under a T7 promoter was ordered from Origene (RC218518), and was sequence verified by Primordium Labs upon receipt.

### Polypeptide pools for activation induced marker (AIM) assays

To obtain polypeptides tiling full length SNX8 protein, 15-mer polypeptide fragments with 11 amino acid overlaps were ordered from JPT Peptide Technologies and synthesized. Together a pool of 130 of these polypeptides (referred to as “SNX8 Pool”) spanned all known translated SNX8 (the full length 465 amino acid SNX8 protein, as well as a unique region of SNX8 Isoform 3). A separate pool was designed to cover primarily the region of SNX8 with similarity to the SARS-CoV-2 nucleocapsid protein in high resolution (referred to as “high resolution epitope pool”). This pool contained 20 10-mers with 9 amino acid overlaps tiling amino acids 44 through 72 (IVQQVPAPSRMQMPQGNPLLLSHTLQELL) of full length SNX8 protein. The sequence of each of these 150 polypeptides was verified by mass spectrometry and purity was calculated by high-performance liquid chromatography (HPLC).

### Peptides for tetramer assays

For use in loading tetramers, two peptides were ordered from Genemed Synthesis, Inc., as 9-mers. One sequence, LQLPQGITL, corresponds to the region of the SARS-CoV-2 nucleocapsid protein with similarity to human SNX8. This sequence was verified by mass spectrometry and purity was calculated as 96.61% by HPLC. The other sequence, MQMPQGNPL, corresponds to the region of human SNX8 protein with similarity to the SARS-CoV-2 nucleocapsid protein. This sequence was verified by mass spectrometry and purity was calculated as 95.83% by HPLC.

### Human proteome PhIP-Seq

Human Proteome PhIP-Seq was performed following our previously published vacuum-based PhIP-Seq protocol (*45*) (https://www.protocols.io/view/scaled-high-throughput-vacuum-phip-protocol-ewov1459kvr2/v1).

Our human peptidome library consists of a custom-designed phage library of 731,724 unique T7 bacteriophage each presenting a different 49 amino-acid peptide on its surface. Collectively these peptides tile the entire human proteome including all known isoforms (as of 2016) with 25 amino-acid overlaps. 1 milliliter of phage library was incubated with 1 microliter of human serum overnight at 4C and immunoprecipitated with 25 microliters of 1:1 mixed protein A and protein G magnetic beads (Thermo Fisher, Waltham, MA, #10008D and #10009D). These beads were than washed, and the remaining phage-antibody complexes were eluted in 1 milliliter of E.Coli (BLT5403, EMD Millipore, Burlington, MA) at 0.5-0.7 OD and amplified by growing in 37C incubator. This new phage library was then re-incubated with the same individual’s serum and the previously described protocol repeated. DNA was then extracted from the final phage-library, barcoded, and PCR-amplified and Illumina adaptors added. Next-Generation Sequencing was then performed using an Illumina sequencer (Illumina, San Diego, CA) to a read depth of approximately 1 million per sample.

### Human proteome PhIP-Seq analysis

All human peptidome analysis (except when specifically stated otherwise) was performed at the gene-level, in which all reads for all peptides mapping to the same gene were summed, and 0.5 reads were added to each gene to allow inclusion of genes with zero reads in mathematical analyses. Within each individual sample, reads were normalized by converting to the percentage of total reads. To normalize each sample against background non-specific binding, a fold-change (FC) over mock-IP was calculated by dividing the sample read percentage for each gene by the mean read-percentage of the same gene for the AG bead only controls. This FC signal was then used for side-by-side comparison between samples and cohorts. FC values were also used to calculate z-scores for each MIS-C patients relative to controls and for each control sample by using all remaining controls. These z-scores were used for the logistic-regression feature weighting. In instances of peptide-level analysis, raw reads were normalized by calculating the number of reads per 100,000 reads.

### SARS-CoV-2 proteome PhIP-Seq

SARS-CoV-2 Proteome PhIP-Seq was performed as previously described(*51*). Briefly, 38 amino acids fragments tiling all open reading frames from SARS-CoV-2, SARS-CoV-1, and 7 other CoVs were expressed on T7 bacteriophage with 19 amino acid overlaps. 1 milliliter of phage library was incubated with 1 microliter of human serum overnight at 4C and immunoprecipitated with 25 microliters of 1:1 mixed protein A and protein G magnetic beads (Thermo Fisher, Waltham, MA, #10008D and #10009D). Beads were washed 5 times on a magenetic plate using a P1000 multichannel pipette. The remaining phage-antibody complexes were eluted in 1 milliliter of E.Coli (BLT5403, EMD Millipore, Burlington, MA) at 0.5-0.7 OD and amplified by growing in 37C incubator. This new phage library was then re-incubated with the same individual’s serum and the previously described protocol repeated for a total of 3 rounds of immunoprecipitations. DNA was then extracted from the final phage-library, barcoded, and PCR-amplified and Illumina adaptors added. Next-Generation Sequencing was then performed using an Illumina sequencer (Illumina, San Diego, CA) to a read depth of approximately 1 million per sample.

### Coronavirus proteome PhIP-Seq analysis

To account for differing read depths between samples, the total number of reads for each peptide fragment was converted to the number of reads per 100k (RPK). To calculate normalized enrichment relative to pre-COVID controls (FC > Pre-COVID), the RPK for each peptide fragment within each sample was divided by the mean RPK of each peptide fragment among all pre-COVID controls. These FC > Pre-COVID values were used for all subsequent analyses as described in the text and figures.

### Radioligand binding assay (RLBA)

Radioligand binding assays were performed as previously described(*45, 46*).Briefly, DNA plasmids containing full-length cDNA under the control of a T7 promoter for each of the validated antigens (see RLBA plasmids above) were verified by Primordium Labs sequencing. The respective DNA templates were used in the T7 TNT in vitro transcription/translation kit (Promega, Madison, WI; #L1170) using [35S]-methionine (PerkinElmer, Waltham, MA; #NEG709A). Respective protein was column-purified on Nap-5 columns (GE healthcare, Chicago, IL; #17-0853-01), and equal amounts of protein (approximately 35,000 counts per minute (cpm)) were incubated overnight at 4 degrees C with 2.5 ul of serum or 1 ul anti-Myc positive control antibody (Cell Signaling Technology, #2272 S). Immunoprecipitation was then performed on 25 microliters of Sephadex protein A/G beads (Sigma Aldrich, St. Louis, MO; #GE17-5280-02 and #GE17-0618-05, 4:1 ratio) in 96-well polyvinylidene difluoride filtration plates (Corning, Corning, NY; #EK-680860). After thoroughly washing, the counts per minute (cpm) of immunoprecipitated protein was quantified using a 96-well Microbeta Trilux liquid scintillation plate reader (Perkin Elmer).

### Split luciferase binding assay (SLBA)

SLBA was performed as described in Rackaityte et. al(*61*). A detailed SLBA protocol is available on protocols.io at: dx.doi.org/10.17504/protocols.io.4r3l27b9pg1y/v1.

Briefly, the DNA oligomers listed above (SLBA DNA oligomers) were amplified by PCR using the primer pairs listed above (SLBA DNA oligomers). Unpurified PCR product was used as input in the T7 TNT in vitro transcription/translation kit (Promega, Madison, WI; #L1170) and Nano-Glo HiBit Lytic Detection System (Promega Cat No. N3040) was used to measure relative luciferase units (RLU) of translated peptides in a luminometer. Equal amounts of protein (in the range of 2e6 to 2e7 RLU) were incubated overnight with 2.5 ul patient sera or 1ul anti-HiBit positive control antibody (Promega, Madison, WI; #CS2006A01)at 4 C. Immunoprecipitation was then performed on 25 microliters of Sephadex protein A/G beads (Sigma Aldrich, St. Louis, MO; #GE17-5280-02 and #GE17-0618-05, 1:1 ratio) in 96-well polyvinylidene difluoride filtration plates (Corning, Corning, NY; #EK-680860). After thoroughly washing, luminescence was measured using Nano-Glo HiBit Lytic Detection System (Promega Cat No. N3040) in a luminometer.

### Activation induced marker (AIM) assay

Peripheral blood mononuclear cells (PBMCs) were obtained from 10 patients with MIS-C and 10 controls for use in the AIM assay. PBMCs were thawed, washed, resuspended in serum-free RPMI medium, and plated at a concentration of 1e10^6^ cell/well in a 96-well round-bottom plate. For each individual, PBMCs were stimulated for 24-hours with either the SNX8 pool (see above) at a final concentration of 1 ug/mL/peptide in 0.2% DMSO, or a vehicle control containing 0.2% DMSO only. For 4 of the controls and 2 of the MIS-C patients, there were sufficient PBMCs for an additional stimulation condition using the SNX8 high resolution epitope pool (see above) also at a concentration of 1 ug/mL/peptide in 0.2% DMSO for 24-hours. Following the stimulation, cells were washed with FACS buffer (Dulbecco’s PBS without calcium or magnesium, 0.1% sodium azide, 2 mM EDTA, 1% FBS) and stained with the following antibody panel for 20 minutes at 4 degrees and then flow cytometry analysis was immediately performed:

| Antibody Target | Clone | Fluorophore | Vendor | Catalog # |
| --- | --- | --- | --- | --- |
| CD3 | OKT3 | Alexa 647 | BioLegend | 317312 |
| CD4 | OKT4 | Alexa 488 | BioLegend | 317420 |
| CD8 | SK1 | Alexa 700 | BioLegend | 344724 |
| OX-40 (CD134) | ACT35 | PE-Dazzle 594 | BioLegend | 350020 |
| CD69 | FN-50 | PE | BioLegend | 310906 |
| CD137 (4-1BB) | 4B4-1 | BV421 | BioLegend | 309820 |
| CD14 | HCD14 | PerCP-Cy5.5 | BioLegend | 325622 |
| CD16 | B73.1 | PerCP-Cy5.5 | BioLegend | 360712 |
| CD19 | HIB19 | PerCP-Cy5.5 | BioLegend | 302230 |
| Live/dead dye |  | eFluor 506 | Invitrogen | 65-0866-14 |

The AIM analysis was performed using FlowJo software using the gating strategy shown in Supplemental Figure 10A. All gates were fixed within each condition of each sample. Activated CD4 T-cells were defined as those which were co-positive for OX40 and CD137. Activated CD8 T-cells were defined as those which were co-positive for CD69 and CD137. Gating thresholds for activation were defined by the outer limits of signal in the vehicle controls allowing for up to 2 outlier cells. Frequencies were calculated as a percentage of total CD3 positive cells (T-cells). Two MIS-C samples had insufficient total events captured by flow cytometry (total of 5,099 and 4,919 events, respectively) and were therefore removed from analysis.

### Tetramer Assay

PBMCs from 2 MIS-C patients with HLA-A*02.01 (1 identified by PAXGene genotyping, 1 by serotyping) and 1 MIS-C patient with HLA-B*35.01 (identified by PAXGene genotyping), and 3 at-risk controls with HLA-A*02.01 (identified by serotyping) were thawed, washed, and put into culture with media containing recombinant human IL-2 at 10 ng/mL in 96-well plates. Peptide fragments (details above) LQLPQGITL and MQMPQGNPL were then added to PBMCs to a final concentration of 10 ug/mL/peptide and incubated (37C, 5% CO2) for 7 days.

Following the 7 days of incubation, a total of 8 pMHCI tetramers were generated from UV-photolabile biotinylated monomers, 4 each from HLA-A*02:01 and HLA-B*35:01 (NIH Tetramer Core). Peptides were loaded via UV peptide exchange. Tetramerization was carried out using streptavidin conjugated to fluorophores PE and APC or BV421 followed by quenching with 500uM D-biotin similar to our previously published methods(*62, 63*). Tetramers were then pooled together as shown below:

**HLA-A\*02:01 Pool**
| Protein | Epitope | MHC I restriction | Fluorophore |
| --- | --- | --- | --- |
| SARS-CoV-2 NP | LQLPQGITL | A*02:01 | PE |
| SARS-CoV-2 NP | LQLPQGITL | A*02:01 | APC |
| SNX8 | MQMPQGNPL | A*02:01 | PE |
| SNX8 | MQMPQGNPL | A*02:01 | BV421 |

**HLA-B\*35:01 Pool**
| Protein | Epitope | MHC I restriction | Fluorophore |
| --- | --- | --- | --- |
| SARS-CoV-2 NP | LQLPQGITL | B*35:01 | PE |
| SARS-CoV-2 NP | LQLPQGITL | B*35:01 | APC |
| SNX8 | MQMPQGNPL | B*35:01 | PE |
| SNX8 | MQMPQGNPL | B*35:01 | BV421 |

All PBMCs were then treated with 100 nM Dasatinib (StemCell) for 30 min at 37 °C followed by staining (no wash step) with the respective tetramer pool corresponding to their HLA restriction (final concentration, 2 to 3 µg/ml) for 30 min at room temperature. Cells were then stained with the following cell surface markers for 20 minutes, followed by immediate analysis on a flow cytometer:

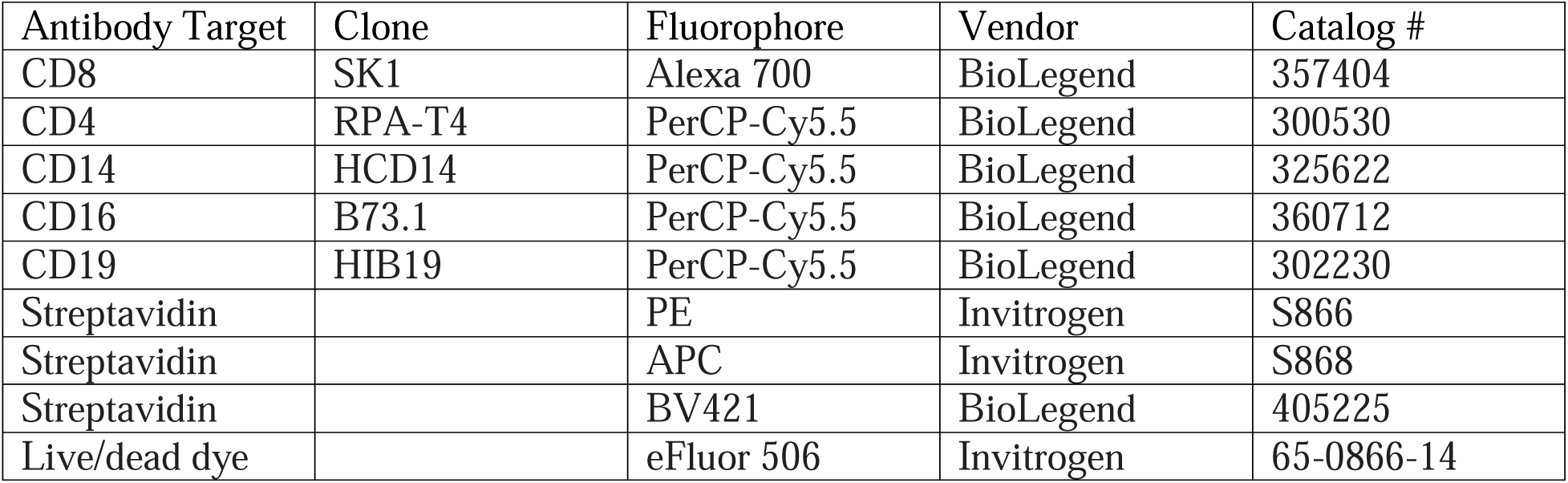

The gating strategy is outlined in Supplemental Figure 10B. A stringent tetramer gating strategy was used to identify cross-reactive T-cells, whereby CD8+ T-cells were required to be triple-positive for PE, APC, and BV421 labels (i.e. a single CD8 T-cell bound to PE conjugated LQLPQGITL and/or PE conjugated MQMPQGNPL in addition to APC-conjugated LQLPQGITL and BV421 conjugated MQMPQGNPL).

Serotyping was performed using an anti-HLA-A2 antibody (FITC anti-human HLA-A2 Antibody, Clone BB7.2, BioLegend, Cat#343303), and pertinent results are shown in Supplemental Figure 10C.

### Single Cell RNA Sequencing Analysis

To assess the cell-type specificity in a relevant disease context, we analyzed SNX8 expression from a single-cell sequencing of PBMC samples from patients with severe, mild, or asymptomatic COVID-19 infection, Influenza infection and healthy controls(*57*). Gene expression data from 59,572 pre-filtered cells was downloaded from the GEO database under accession <u>GSE149689</u> for analysis and downstream processing with scanpy(*64*). Cells with (i) less than 1000 total counts, (ii) less than 800 expressed genes and (iii) more than 3000 expressed genes were filtered out as further quality control, leaving 42,904 cells for downstream analysis. Gene expression data were normalized to have 10,000 counts per cell and were log_1_ *P* transformed. Highly variable genes were calculated using the scanpy function highly_variable_genes using Seurat flavor with the default parameters (min_mean = 0.0125, max_mean = 3, and min_disp = 0.5)(*65*). Only highly variable genes were used for further analysis. The total number of counts per cell was regressed out, and the gene expression matrix was scaled using the scanpy function scale with max_value = 10. Dimensionality reduction was performed using principal components analysis with 50 principal components. Batch balanced kNN (k-nearest neighbors), implemented with scanpy’s function bbknn, was used to compute each cell’s top neighbors and normalize batch effects(*66*). The batch-corrected cells were clustered using the Leiden algorithm and projected into two dimensions with uniform manifold approximation and projection (UMAP) for visualization. Initial cluster identity was determined by finding marker genes with differential expression analysis preformed using a *t* test on log_1_ *P*– transformed raw counts with the scanpy function rank_genes_groups(*67, 68*).

### Statistical methods

All statistical analysis was performed in Python using the Scipy Stats package. For comparisons of distributions of PhIP-Seq enrichment between two groups, a non-parametric Kolmogorov-Smirnov test was utilized. For logistic regression feature weighting, the Scikit-learn package(*69*) was used, and logistic regression classifiers were applied to z-scored PhIP-Seq values from individuals with MIS-C versus at-risk controls. A liblinear solver was used with L1 regularization, and the model was evaluated using a five-fold cross-validation (4 of the 5 for training, 1 of the 5 for testing). For RLBAs and SLBAs, first an antibody index was calculated as follows: (sample value – mean blank value) / (positive control antibody values – mean blank values). For the alanine mutagenesis scans, blank values of each construct were combined, and a single mean was calculated. A normalization function was then applied to the experimental samples only (excluding antibody only controls) to create a normalized antibody index ranging from 0 to 1. Comparisons between two groups of samples were performed using a Mann-Whitney U test. An antibody was considered to be “positive” when the normalized antibody index in a sample was greater than 3 standard deviations above the mean of controls. When comparing two groups of normally distributed data, a T-test was performed. P-values: ns > 0.05, * < 0.05, ** < 0.01, *** < 0.001, **** < 0.0001, ***** < 0.00001.

## Supplemental Tables

**Supplemental Table 1:** Phenotypic data for MIS-C patients.

| **Organ System Involvement** (number reporting) | **Phenotype Present** | **Percent** |
| --- | --- | --- |
| **Respiratory** |  |  |
| Oxygen requirement (198) | 139 | 70.2 |
| Bronchospasm (161) | 2 | 1.2 |
| Acute Respiratory Distress Syndrome (ARDS) (198) | 24 | 12.1 |
| **Cardiac** |  |  |
| Vasoactive support (198) | 124 | 62.6 |
| Myocarditis (199) | 115 | 57.8 |
| Elevated Troponin (197) | 128 | 65 |
| **Neurologic** |  |  |
| Any neurologic symptoms (197) | 22 | 11.2 |
| Encephalitis (197) | 2 | 1 |
| **Gastrointestinal** (196) | 66 | 33.7 |
| **Hematologic** (197) | 22 | 11.2 |
| **Rheumatologic** |  |  |
| Arthritis (195) | 9 | 4.6 |
| Myositis (194) | 27 | 12.9 |
| **Mucocutaneous** |  |  |
| Conjunctivitis (197) | 105 | 53.3 |
| Oral Mucosal Changes (198) | 76 | 38.4 |
| Rash (197) | 106 | 53.8 |

### Table Definitions

Oxygen requirement; receipt of any oxygen support at any time during hospitalization. Bronchospasm; severe bronchospasm requiring continuous bronchodilators. Acute Respiratory Distress Syndrome (ARDS); onset of hypoxemia was acute (during this illness), chest imaging findings of new infiltrates (unilateral or bilateral), respiratory failure not fully explained by cardiac failure or fluid overload, PaO2/FiO2 ratio < 300 or SpO2/FiO2 < 264 (if SpO2 < 97), on CPAP > 5 cm H2O or BiPAP or Invasive Mechanical Ventilation. Vasoactive support; receipt of vasoactive infusions (at any time during hospitalization) including: Dopamine, Dobutamine, Epinephrine, Norepinephrine, Phenylephrine, Milrinone, Vasopressin (for hypotension, not diabetes insipidus). Myocarditis; myocarditis diagnosed during hospital stay and adjudicated by outside panel of cardiologists. Elevated troponin; based on site-specific cutoff. Any neurologic symptoms; suspected central nervous system infection, stroke or intracranial hemorrhage (at presentation or during hospitalization), seizure (at presentation or during hospitalization), coma or unresponsive, receipt of neurodiagnostic imaging (CT, MRI, or LP), encephalitis, decreased hearing, decreased vision, iritis or uveitis. Gastrointestinal; appendicitis, diarrhea (at presentation or during hospitalization), abdominal pain (at presentation or during hospitalization), gallbladder hydrops or edema, pancreatitis, hepatitis, nausea/loss of appetite at presentation, vomiting at presentation. Hematologic; anemia with hemoglobin <9 g/dL, minimum white blood cells <4 x 10^3^ cells/µL, minimum platelets <150 x 10^3^ cells/µL, deep vein thrombosis, pulmonary embolism, hemolysis, bleeding, ischemia of an extremity. Oral Mucosal Changes; erythema of lips or oropharynx, strawberry tongue, or drying or fissuring of the lips.

**Supplemental Table 2:** Clinical characteristics of validation cohorts

|  | Severe Acute COVID-19 | MIS-C Validation |
| --- | --- | --- |
| Number | 29 | 24 |
| Male sex (%) | 14 (48.3) | 16 (66.7) |
| Median age in years (median, IQR) | 11.4 (11.4) | 7.7 (8.6) |
| **Race and ethnicity (%)** |  |  |
| White, non-Hispanic | 14 (48.3) | 3 (12.5) |
| Black, non-Hispanic | 4 (13.8) | 8 (33.3) |
| Hispanic or Latino | 9 (31.0) | 13 (54.2) |
| Other race, non-Hispanic | 2 (6.9) | 0 (0.0) |
| **Underlying conditions (%)** |  |  |
| None | 9 (31.0) | 13 (54.2%) |
| Obesity* | 8 (27.6) | 7 (29.2) |
| Asthma | 3 (10.3) | 2 (8.3) |
| Cardiovascular | 1 (3.4) | 0 (0.0) |
| Immunocompromise | 1 (3.4) | 0 (0.0) |
| Autoimmune condition | 0 (0.0) | 0 (0.0) |
| Malignancy | 1 (3.4) | 0 (0.0) |
| **Interventions (%)** |  |  |
| ICU admission | 22 (75.9) | 11 (45.8) |
| Shock requiring vasopressors | 11 (37.9) | 0 (0.0) |
| Mechanical ventilation | 6 (20.7) | 2 (8.3) |

\*Does not include n=4 acute COVID-19 patients and n=1 MIS-C patients under 2 years of age.

## Data Availability

All data produced in the present study are available upon reasonable request to the authors. Complete data will be made available via a Dryad repository upon peer-reviewed publication: doi:10.7272/Q6SJ1HVH

## Acknowledgements

The following members of the Overcoming COVID-19 Network Study Group Investigators (presented alphabetically by state) were all closely involved with the design, implementation, and oversight of the Overcoming COVID-19 study as well as collecting patient samples and data. *Alabama:* Children’s of Alabama, Birmingham: Michele Kong, MD, Heather Kelley, RN, BSN; Meghan Murdock, RN, BSN; Candice Colston. *Arizona*: Katri V. Typpo, MD. *Arkansas:* Arkansas Children’s Hospital, Little Rock: Katherine Irby, MD; Ronald C. Sanders Jr, MD, MS; Masson Yates; Chelsea Smith. *California:* UCSF Benioff Children’s Hospital Oakland, Oakland: Natalie Z. Cvijanovich, MD; UCSF Benioff Children’s Hospital, San Francisco: Matt S. Zinter, MD. *Colorado:* Aline B. Maddux, MD, MSCS, Emily Port, BA, PMP; Rachel Mansour, BSN, RN, CPN; Sara Shankman, DNP, CPNC-AC, Natasha Baig, MBBS, Frances Zorensky, BS.. *Florida:* Holtz Children’s Hospital, Miami: Paula S. Espinal, MD, MPH; Brandon Chatani, MD; Gwenn McLaughlin, MD, MSPH. *Georgia:* Children’s Healthcare of Atlanta at Egleston, Atlanta: Keiko M. Tarquinio, MD; Kaitlin Jones, MSN, RN, CCRP. *Illinois:* Ann & Robert H. Lurie Children’s Hospital of Chicago, Chicago: Bria M. Coates, MD. *Indiana:* Riley Children’s Hospital, Indianapolis: Courtney M. Rowan, MD, MScr. *Massachusetts:* Boston Children’s Hospital, Boston: Adrienne G. Randolph, MD; Margaret M. Newhams, MPH; Suden Kucukak, MD; Tanya Novak, PhD; Elizabeth R. McNamara BSN, RN; Hye Kyung Moon, MA; Takuma Kobayashi BS; Jeni Melo, BS; Sergio R. Jackson, MS; Marah Kiana Echon Rosales, BS; Cameron Young, BS; Sabrina R. Chen, BS; Janet Chou, MD; Rezende Da Costa Aguiar, PhD; Maria Gutierrez-Arcelus, PhD, Megan Elkins, MHS. Taking On COVID-19 Together team: David Williams, MD; Lucinda Williams, DNP, MSN, RN, PNP, NE-BC; Leah Cheng, MA; Yubo Zhang, BS; Danielle Crethers, BA; Debra Morley, PhD; Sarah Steltz, MPH; Kelly Zakar, MSN, RN, PPCNP-BC; Myriam A. Armant, PhD; Felicia Ciuculescu, MD; *Michigan:* University of Michigan C. S. Mott Children’s Hospital, Ann Arbor: Heidi R. Flori, MD, FAAP; Mary K. Dahmer, PhD. *Minnesota:* Mayo Clinic, Rochester: Emily R. Levy, MD, FAAP; Supriya Behl, MSc; Noelle M. Drapeau, BA. *Mississippi:* Charlotte V. Hobbs, MD *Missouri:* Children’s Mercy Hospital, Kansas City: Jennifer E. Schuster, MD; Abigail Kietzman BS; Shannon Hill, BSN. *Nebraska:* Children’s Hospital & Medical Center, Omaha: Melissa L. Cullimore, MD, PhD; Russell J. McCulloh, MD. *New Jersey:* Cooperman Barnabas Medical Center, Livingston: Shira J. Gertz, MD. *North Carolina:* University of North Carolina, Chapel Hill: Stephanie P. Schwartz, MD; Tracie C. Walker, MD. *Ohio:* Akron Children’s Hospital, Akron: Ryan A. Nofziger, MD; Cincinnati Children’s Hospital, Cincinnati: Mary Allen Staat, MD, MPH; Chelsea C. Rohlfs, BS, MBA. *Pennsylvania:* Children’s Hospital of Philadelphia, Philadelphia: Julie C. Fitzgerald, MD, PhD, MSCE; Ryan Burnett, BS, Jenny Bush, RNC, BSN. *South Carolina:* MUSC Shawn Jenkins Children’s Hospital, Charleston: Elizabeth H. Mack, MD, MS; Nelson Reed, MD. *Tennessee:* Monroe Carell Jr Children’s Hospital at Vanderbilt, Nashville: Natasha B. Halasa, MD, MPH. *Texas:* Texas Children’s Hospital and Baylor College of Medicine, Houston: Laura L. Loftis, MD. *Utah:* Primary Children’s Hospital and University of Utah, Salt Lake City: Hillary Crandall, MD, PhD; Kwabena Krow Ampofo, MD.

Members of the US Centers for Disease Control and Prevention COVID-19 Response Team on the Overcoming COVID-19 Study were Laura D. Zambrano, PhD, MPH, Manish M. Patel, MD, MPH, and Angela P. Campbell, MD, MPH.

The authors acknowledge the New York Blood Center for contributing pre-COVID-19 healthy donor blood samples which were used as controls for the SARS-CoV-2 library PhIP-Seq.

The authors acknowledge the contributions of Weston Browne and Sam Pleasure, MD, PhD for their work investigating potential central nervous system specific autoimmunity in MIS-C; Tirtha Kharel, BS, for his help designing Python code used in the analysis; David Blauvelt, MD, for his ideas regarding the application of advanced statistics to PhIP-Seq data analysis.

Biorender (Biorender.com) was used to build graphics for Figure 1A and Figure 4C.

## Disclaimer

The findings and conclusions in this report are those of the authors and do not necessarily represent the views of the US Centers for Disease Control and Prevention.

## Contributions

Conceptualization: A.B, J.J.S.Jr., S.E.V., J.C., A.G.R., M.S.A., J.L.D. Methodology: A.B, J.J.S.Jr., S.E.V., E.R., C.R.Z., A.F.K., J.V.R., J.C., A.G.R., M.S.A., J.L.D. Performed or contributed to experiments: A.B., J.J.S.Jr., S.E.V., A.M., C.Y.W., A.S., J.V.P., D.J.L.Y., H.K., C.M.B. Formal analysis: A.B., J.J.S.Jr., H.S.M., A.F.K. Patient sample and clinical data acquisition: K.Z., T.N., L.D.Z., A.P.C., A.G.R.K.L.M., L.L.L, C.V.H, K.M.T, M.K, J.C.F, P.S.E, T.C.W, S.P.S., H.C., K.I., M.A.S., C.M.R., J.E.S., N.B.H., S.J.G., E.H.M., A.B.M., N.Z.C., M.S.Z. Clinical data curation: T.N., A.G.R. Writing (original draft): A.B., H.M., J.L.D. Writing (review and editing): A.B., J.C., T.N., H.M., L.D.Z., A.P.C, A.G.R., M.R.W., M.S.A., J.L.D. Supervision: J.C., A.G.R., M.S.A., J.L.D.

## Corresponding authors

Correspondence to Joseph L. DeRisi and Mark S. Anderson.

---

**Supplemental Figure 1:**
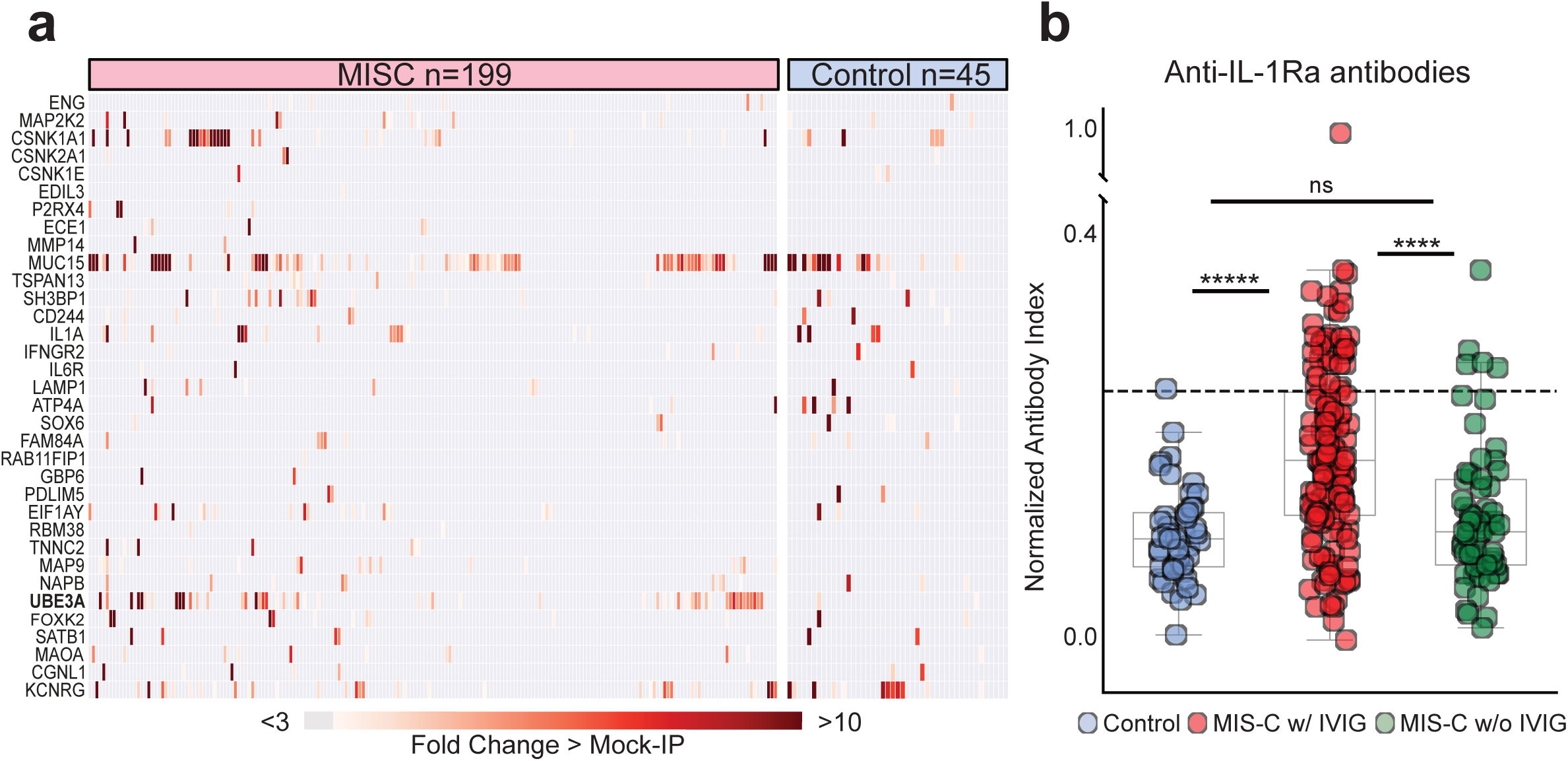
Previously reported MIS-C associated autoantigens. (a) Heatmap showing distribution of PhIP-Seq enrichments (FC > Mock-IP) of previously reported MIS-C autoantibodies in MIS-C patients (n=199) and at-risk controls (n=45). **(b)** Stripplots showing distribution of signal (normalized antibody index) for antibodies targeting IL-1 receptor antagonist (IL-1Ra) measured by RLBA in at-risk controls (blue; n=45), MIS-C patient samples containing IVIG (red; n=135), and MIS-C patient samples without IVIG (green; n=61). Dotted line at 3 standard deviations above the mean of controls. For the figure, Mann-Whitney U testing was performed; ns p-value > 0.05, ****p-value < 0.0001, *****p-value < 0.00001.

**Supplemental Figure 2:**
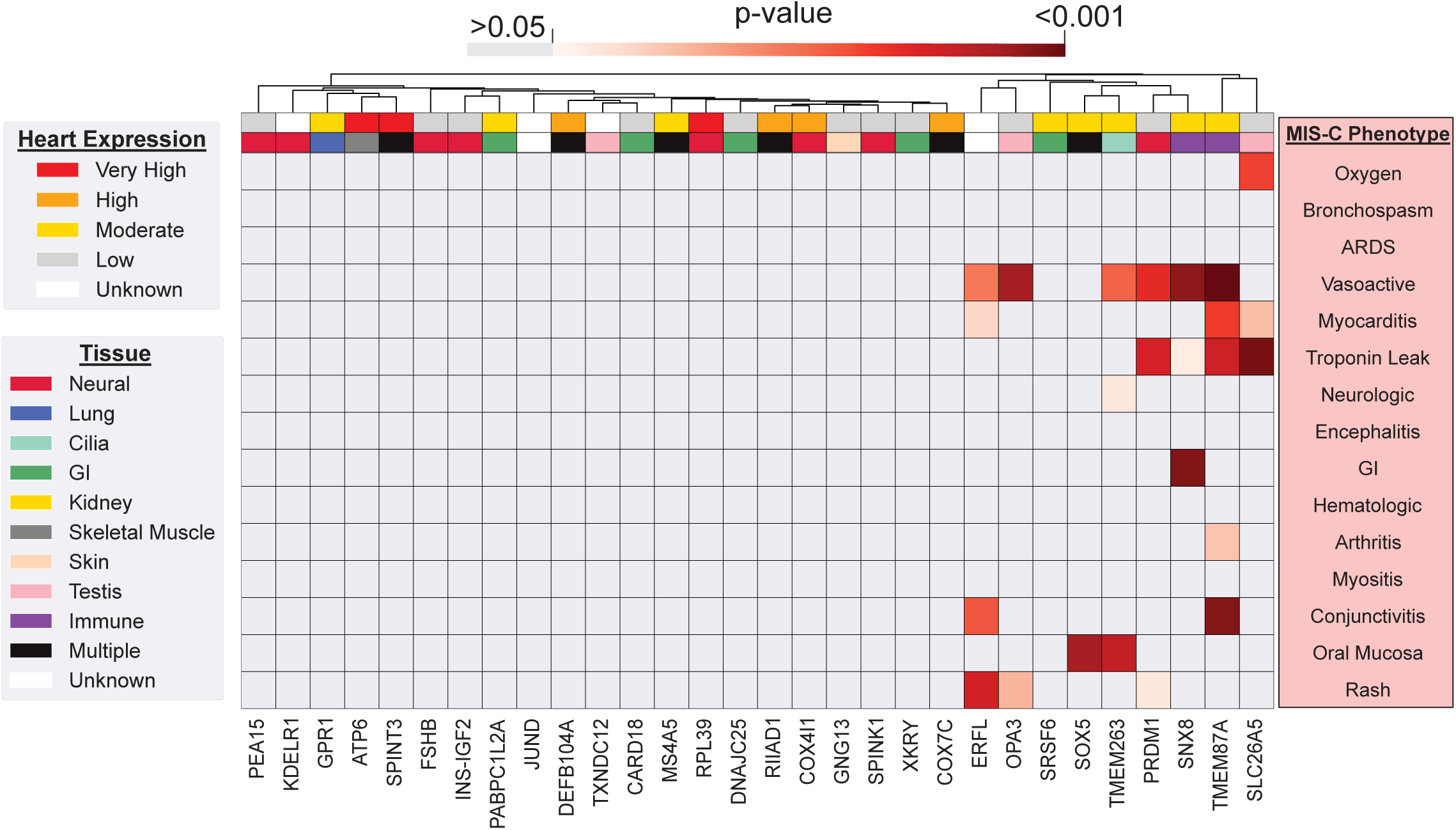
Absence of tissue specific autoantibodies correlating to phenotypes. Heatmap of p-values (Kolmogorov-Smirnov testing) for differences in autoantibody enrichment for MIS-C patients with versus without each clinical phenotype. Significant p-values in the negative direction (in which there is increased signal in individuals without the phenotype) are masked (colored as p > 0.05). For each autoantigen, tissue RNA-sequencing data from Human Protein Atlas (Proteinatlas.org) is shown. Amount of expression in cardiac tissue in top row (Very high=nTPM >1000, High=nTPM 100-1000, Moderate=nTPM 10-100, Low=nTPM <10), and predominant tissue type in second-from-top row. Explanations of criteria for MIS-C phenotypes, and distribution of each phenotype within the cohort, can be found in Supplemental Table 1.

**Supplemental Figure 3:**
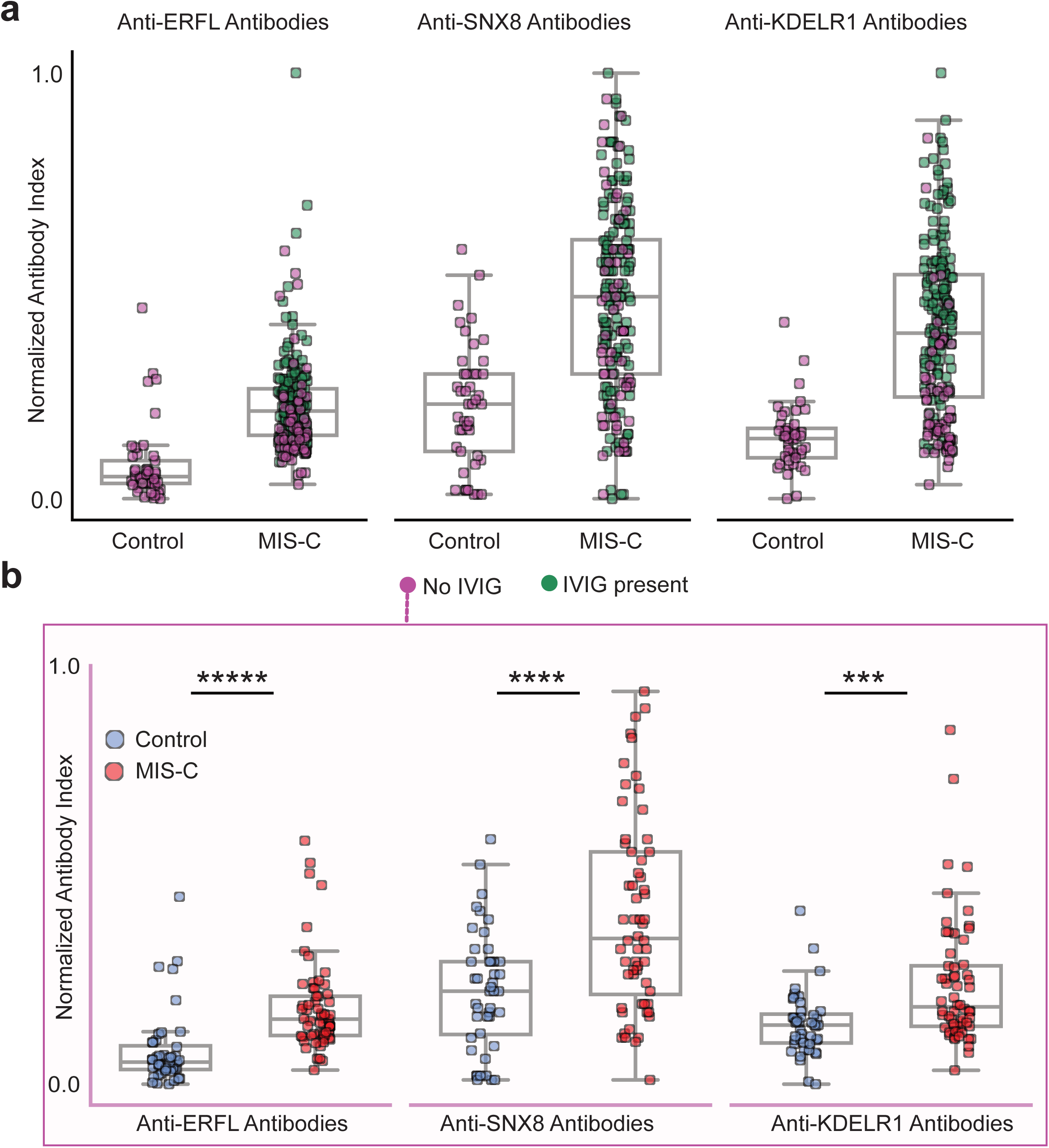
Significantly increased autoantibodies is MIS-C samples without IVIG. (a) Stripplots showing RLBA enrichments (normalized antibody indices) in samples containing IVIG (green) and samples without IVIG (magenta) for ERFL, SNX8, and KDELR1 in MIS-C patients (n=196) and at-risk controls (n=45). **(b)** Stripplots showing RLBA enrichment only in those MIS-C samples without IVIG (n=62) relative to at-risk controls (n=45). For the figure, Mann-Whitney U testing was performed; ***p-value < 0.001, ****p-value < 0.0001, *****p-value < 0.00001.

**Supplemental Figure 4:**
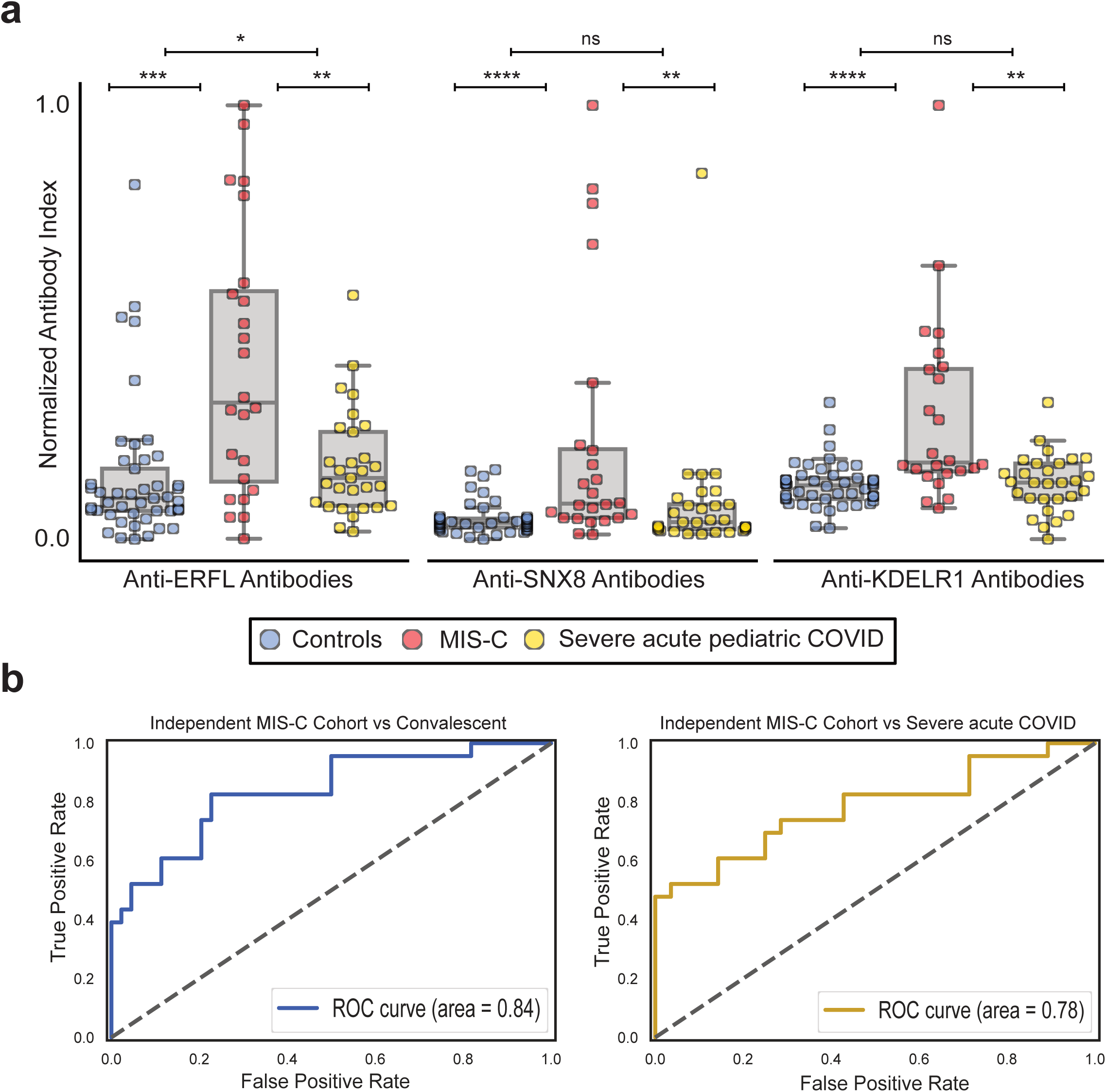
Validation of autoantibody classifier using independent MIS-C cohort. (a) Stripplots showing RLBA enrichments (normalized antibody indices) for ERFL, SNX8, and KDELR1 in an independent cohort of children with MIS-C (red; n=24) compared to children severely ill with acute COVID-19 (yellow; n=29) and at-risk controls (blue; n=45) (**b**) Logistic regression receiver operating characteristic curves for classification of the independent MIS-C cohort versus at-risk controls (left) and the independent MIS-C cohort versus children severely ill with acute COVID-19 (right). For the figure, Mann-Whitney U testing was performed; ns p-value > 0.05, *p-value < 0.05, ** p-value < 0.01, ***p-value < 0.001, ****p-value < 0.0001.

**Supplemental Figure 5:**
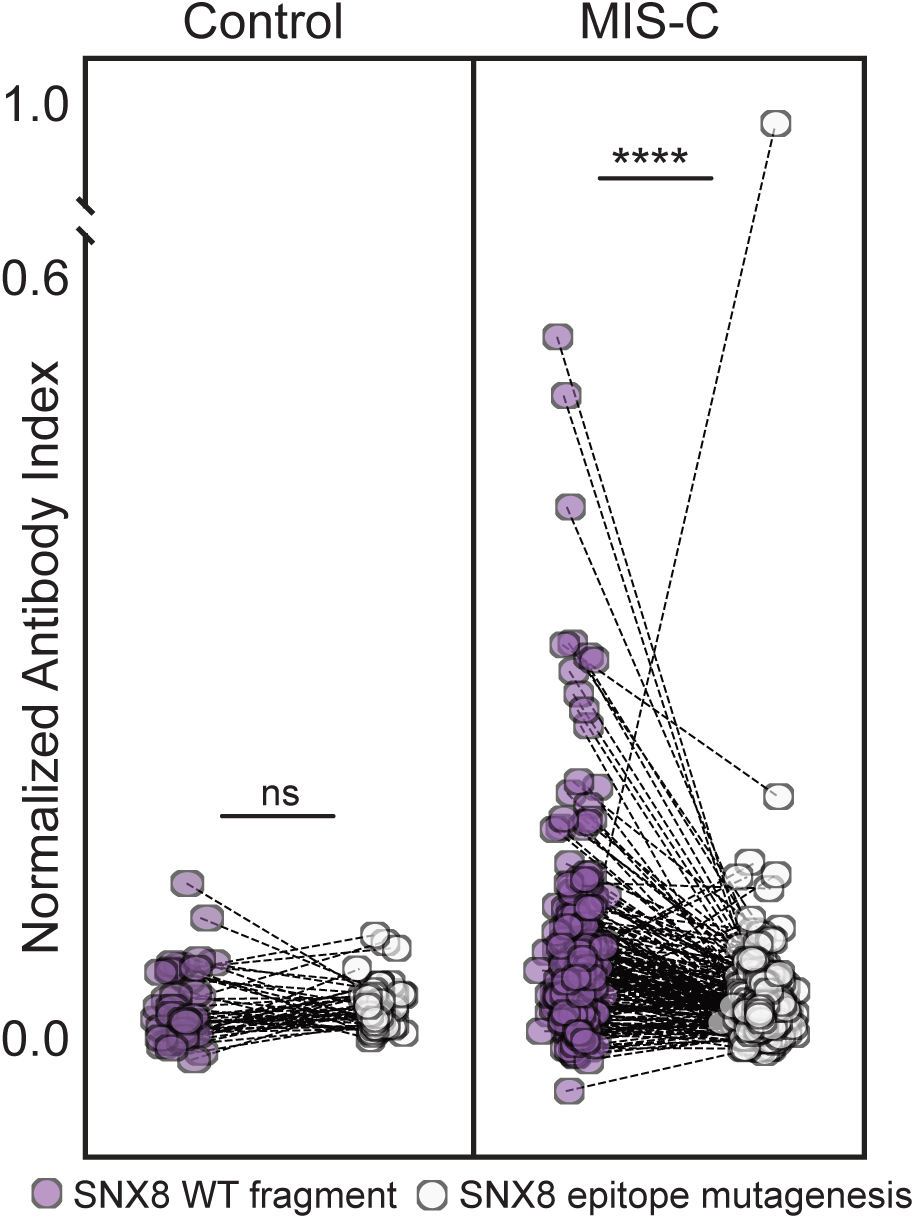
Prevalence of epitope specific SNX8 autoantibodies in MIS-C and controls. Paired stripplots showing SLBA enrichments (normalized antibody indices) in MIS-C patients (n=182) and at-risk controls (n=45) for the full 49 amino acid SNX8 wild-type (WT) polypeptide fragment (lavender) relative to the same SNX8 fragment with alanine mutagenesis of the [PSRMQMPQG] epitope (white). SNX8 WT fragment SLBA values are the means of technical replicates, SNX8 epitope mutagenesis values are from a single experiment. For the figure, Mann-Whitney U testing was performed; ns p-value > 0.05, ****p-value < 0.0001.

**Supplemental Figure 6:**
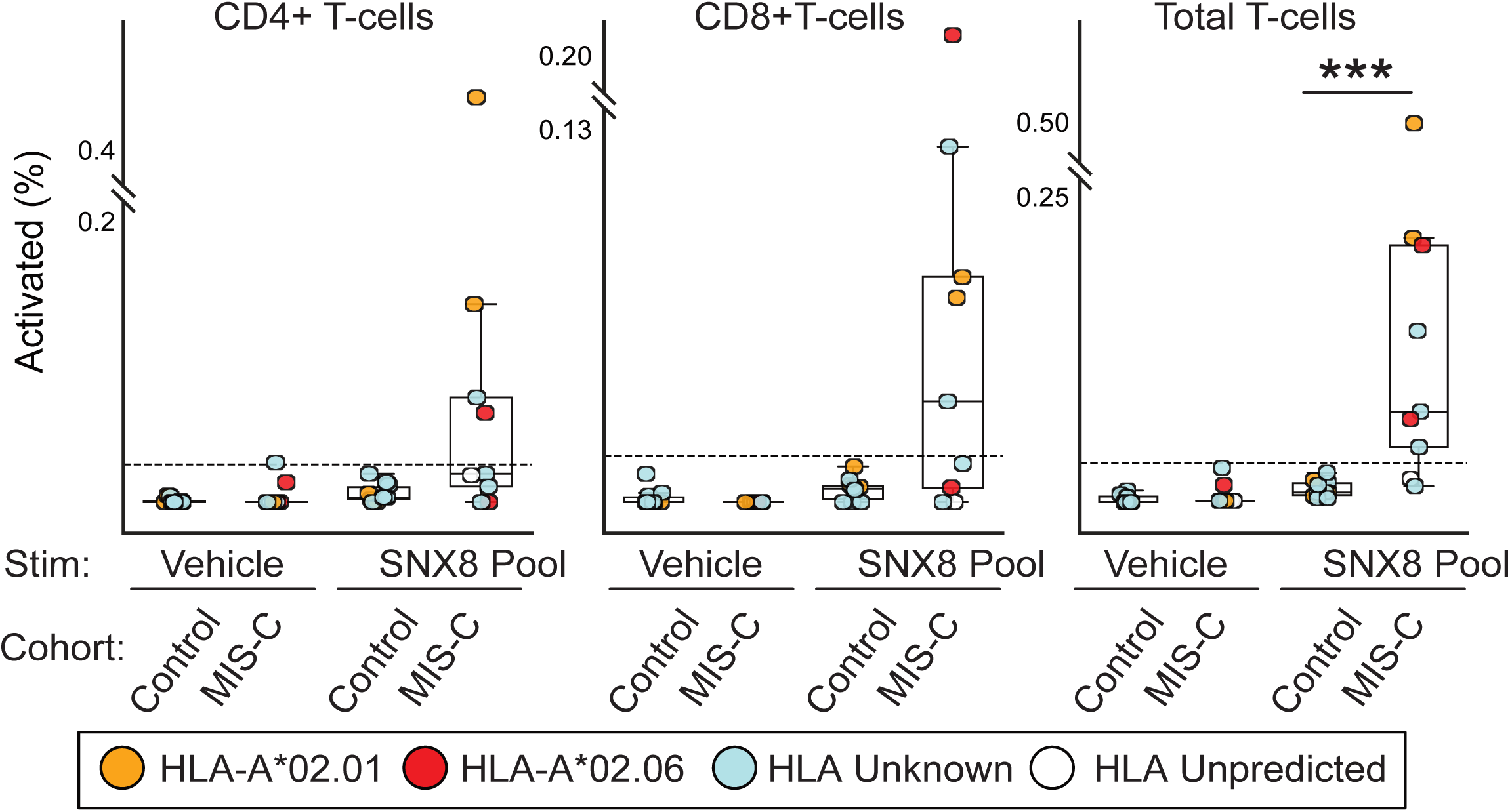
Activation of CD4+ and CD8+ T-cells with associated HLA type. Stripplots showing distribution of CD4+, CD8+, and total T-cells which activate in response to either vehicle (culture media + 0.2% DMSO) or SNX8 peptide pool (SNX8 peptide + culture media + 0.2% DMSO) using AIM assay. HLA type indicated by color of dot. HLA unpredicted means patient contained none of the MIS-C associated HLA types. Dotted line at 3 standard deviations above the mean of the SNX8 stimulated controls. For the figure, Mann-Whitney U testing was performed; ***p-value <0.001.

**Supplemental Figure 7:**
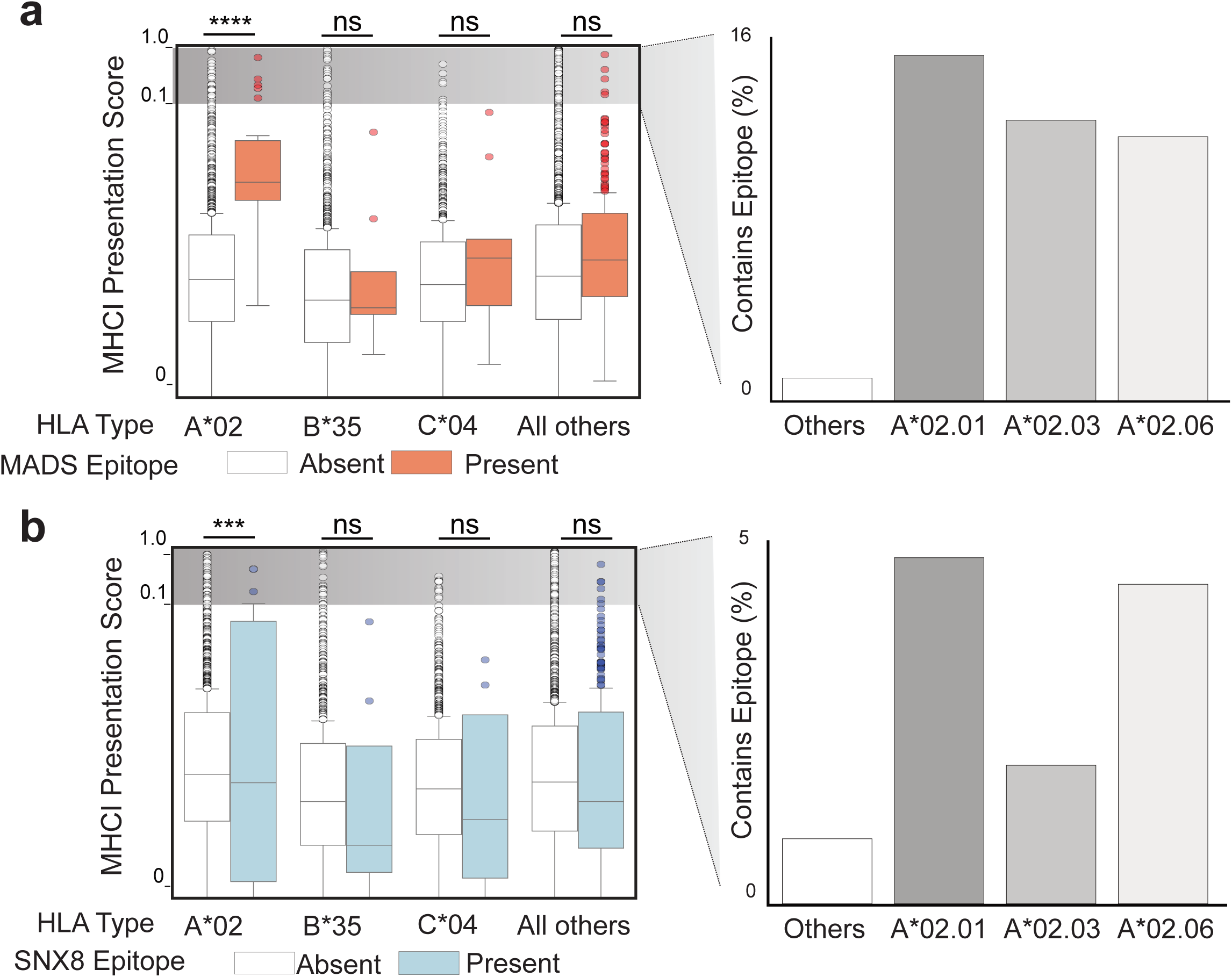
HLA-A*02 preferentially displays MADS and SNX8 similarity regions on MHC1. (a) Computationally predicted MHCI presentation scores (Immune Epitope Database; IEDB.org) for each possible peptide fragment of full length SARS-CoV-2 N protein for each of the three MIS-C associated HLA types (A*02, B*35 and C*04) relative to a reference set of HLA-types encompassing over 99% of humans. Those fragments containing the MADS similarity region “LQLPQG” in orange. Percent of fragments within each specific HLA sub-type with a score greater than 0.1 (likely to be presented) shown on right. **(b)** Identical analysis but using full length SNX8 protein rather than SARS-CoV-2 NP, and the SNX8 similarity region “MGMPQG” rather than the MADS region “LQLPQG”. For the figure, the peptide MHCI presentation scores for all peptides within an HLA type were normally distributed, so a t-test was used to compare signal. ns p-value > 0.05, ***p-value <0.001, ****p-value < 0.0001.

**Supplemental Figure 8:**
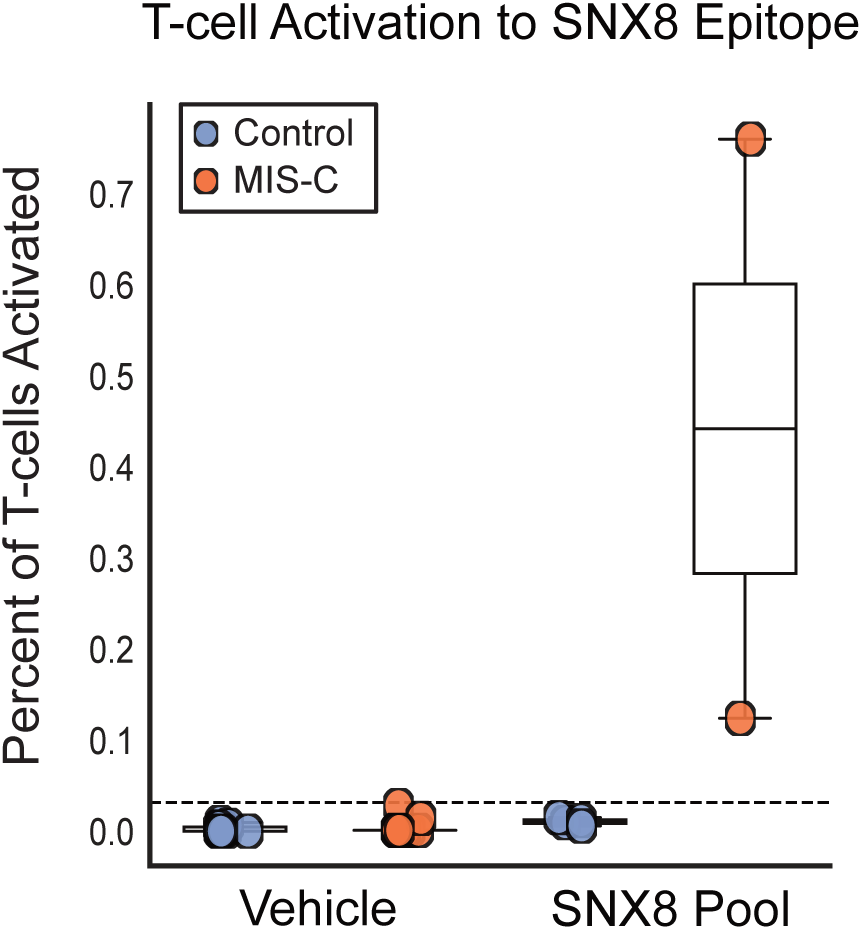
Stimulation with SNX8 epitope only is sufficient to activate T-cells in MIS-C patients. Stripplots showing percentage of total T-cells which activate in response to either vehicle (culture media + 0.2% DMSO) or the SNX8 Epitope (SNX8 Epitope (Materials) + culture media + 0.2% DMSO) in MIS-C patients (n=2) and at-risk controls (n=4) measured by AIM assay. Dotted line at 3 standard deviations above mean of SNX8 Epitope stimulated controls.

**Supplemental Figure 9:**
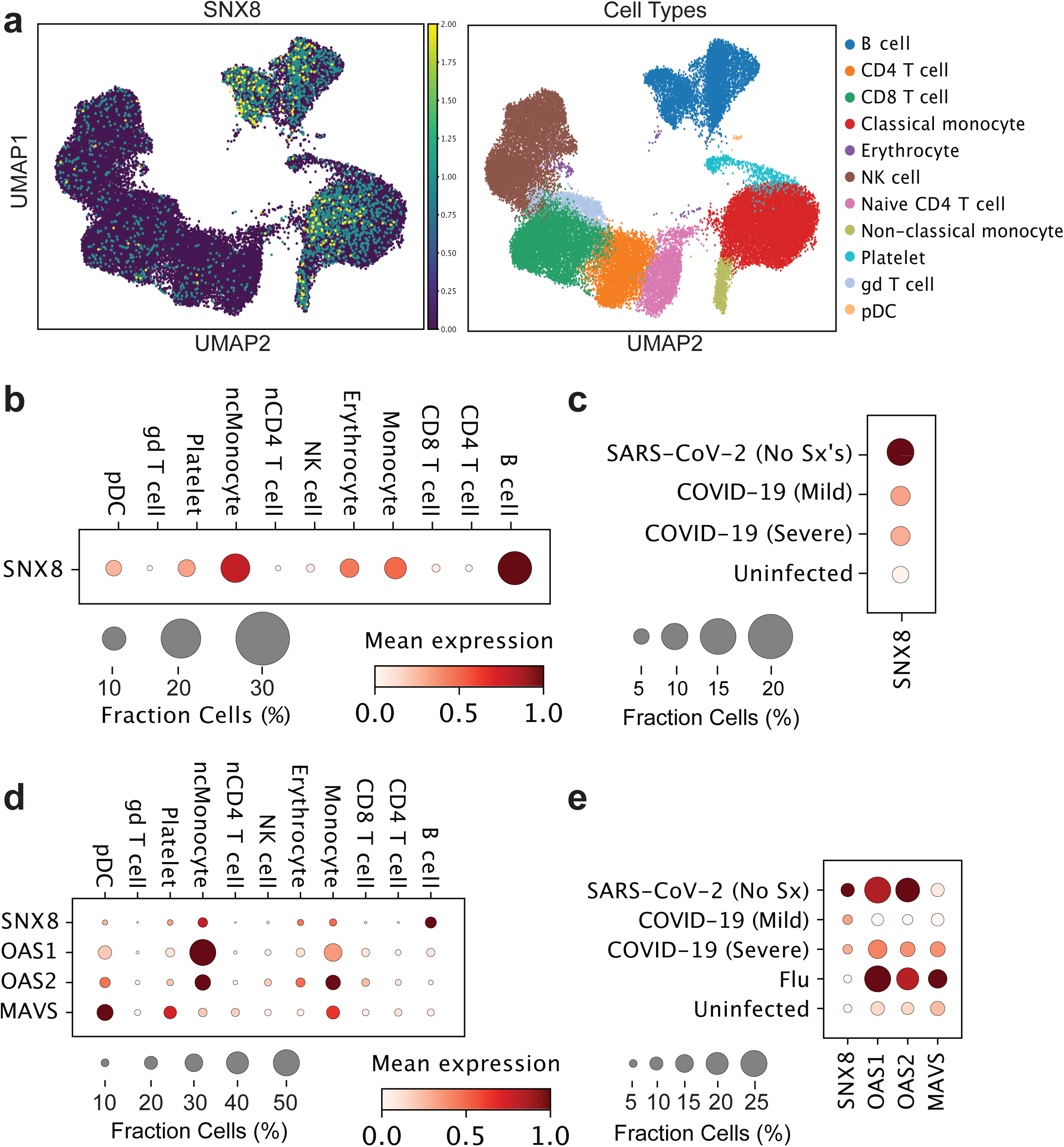
SNX8 expression during viral infection. (**a**) UMAPs showing SNX8 expression in various peripheral blood cell types during SARS-CoV-2 infection. (**b**) Mean expression and percent of cells expressing SNX8 in peripheral blood subsets during SARS-CoV-2 infection. (**c**) Mean expression and percent of cells expressing SNX8 averaged across all peripheral blood mononuclear cells from SARS-CoV-2 infected individuals without symptoms, with mild symptoms, or with severe disease compared to uninfected controls. (**d**) Mean expression and percent of cells expressing SNX8, OAS1, OAS2, and MAVS in peripheral blood subsets during SARS-CoV-2 infection. (**e**) Relative expression of SNX8, OAS1, OAS2, and MAVS during influenza infection compared to different severities of SARS-CoV-2 infection.

**Supplemental Figure 10:**
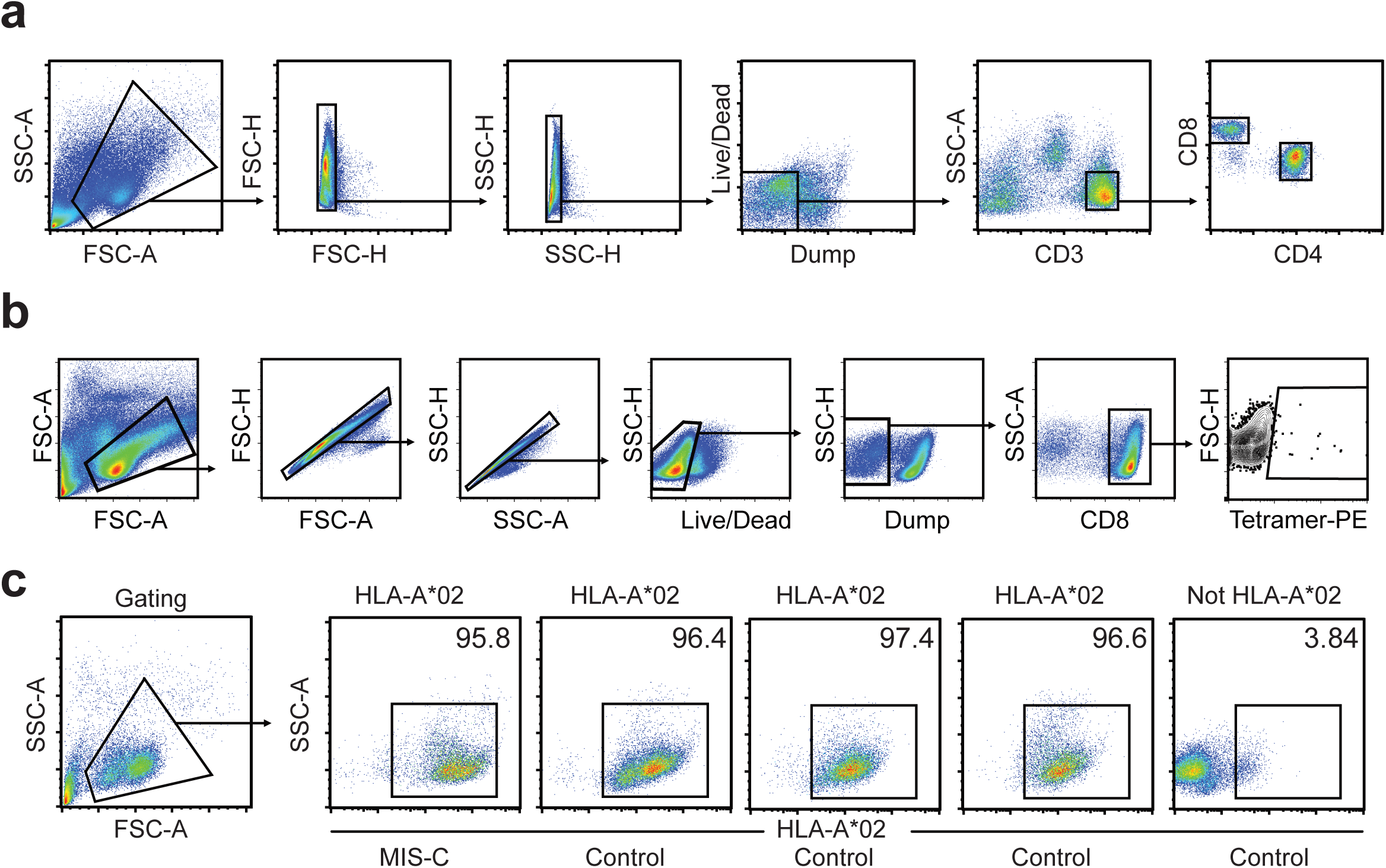
Representative flow cytometry for AIM, tetramer, and serotyping experiments. (a) FlowJo plots showing gating strategy for identifying CD4 positive and CD8 positive T-cells for the AIM analysis shown in Figure 4. **(b)** FlowJo gating strategy for the tetramer assay showing isolation of PE-tetramer positive CD8 positive T-cells. **(c)** FlowJo plots showing results of serotyping for the PBMCs used in the SNX8/MADS tetramer cross-reactivity assay which did not have sufficient cells for genotyping. Shown is the 1 MIS-C patient (far left) and 3 controls (middle 3) which are positive for HLA-A*02 and were used in the tetramer cross-reactivity assay, and one control negative for HLA-A*02 (far right) which was not used.

